# Using the app “Injurymap©” to provide exercise rehabilitation for people with acute lateral ankle sprains seen at the Hospital Emergency Department – a mixed-method pilot study

**DOI:** 10.1101/2022.01.31.22269313

**Authors:** Jonas Bak, Kristian Thorborg, Mikkel Bek Clausen, Finn Elkjær Johannsen, Jeanette Wassar Kirk, Thomas Bandholm

## Abstract

**Background:** Acute lateral ankle sprains (LAS) account for 4-5% of all Emergency Department (ED) visits. Few patients receive the recommended care of exercise rehabilitation. A simple solution is an exercise app for mobile devices, which can deliver tailored and real-time adaptive exercise programs.

**Purpose:** The purpose of this pilot study was to investigate the use and preliminary effect of an app-based exercise program in patients with LAS seen in the Emergency Department at a public hospital.

**Materials and methods:** We used an app that delivers evidence-based exercise rehabilitation for LAS using machine-based learning. Participants were recruited from the ED and followed for four months. Data on app-use and preliminary effect were collected continuously through the exercise app and weekly text-messages. Baseline and follow-up data were collected though an online questionnaire. Semi-structured interviews were performed after participants stopped using the app.

**Results:** Health care professionals provided 485 patients with study information and exercise equipment. Of those, 60 participants chose to enroll in the study and 43 became active users. The active users completed a median of 7 exercise sessions. Most of the active users were very satisfied or satisfied (79%-93%) with the app and 95.7% would recommend it to others. The interviews showed that ankle sprains were considered an innocuous injury that would recover by itself. Several app users expressed they felt insufficiently informed from the ED health care professionals. Only 39% felt recovered when they stopped exercising, and 33% experienced a recurrent sprain in the study period.

**Conclusion:** In this study, only few patients with LAS became active app users after receiving information in the ED about a free app-based rehabilitation program. We speculate the reason for this could be the perception that LAS is an innocuous injury. Most of the patients starting training were satisfied with the app, although few completed enough exercise sessions to realistically impact clinical recovery. Interestingly more than half of the participants did not feel fully recovered when they stopped exercising and one third experienced a recurrent sprain.

**Trial-identifiers:** https://clinicaltrials.gov/ct2/show/NCT03550274, preprint (open access): doi/xxx.

## Introduction

An ankle sprain is one of the most common musculoskeletal injuries with a comprehensive burden for individuals as well as society (1). They account for 4-5% of all Emergency Department visits in Denmark (2) which is consistent with data from other countries (1). This might just be the tip of the iceberg since less than half of the people who sustain an ankle sprain seek health care (1,3). A lateral ankle sprain (LAS) is often regarded as an innocuous injury (4) and especially health care professionals tend to overestimate the recovery (5). However, 32-74% of people who sustain a LAS have prolonged symptoms such as pain, decreased function and subjective instability for several years after their initial injury (1,6). In sport, up to 34% will sustain a recurrent sprain in the following years after their initial injury (6). However, exercise therapy is a well-documented cost-effective rehabilitation modality to treat LAS (7–10) and prevent recurrent sprains(8,11,12). It is unfortunate that few patients are prescribed exercise programs or physiotherapy after a LAS, and that most expenses relate to diagnostic procedures rather than exercise-based rehabilitation (4). Technologically supported self-management may be a solution to this problem but requires investigation.

Applications for smart devices (apps) have the potential to be powerful tools in providing easily accessible exercise therapy programs and in attaining important information about exercise behavior that have been practically unobtainable previously (13). They represent a flexible telehealth solution that most often does not require the online presence of a health care professional. Apps have the ability through interaction between users and smartphone to tailor specific information and exercise programs. Furthermore, apps can give real-time, real-life feedback during a exercise session without waiting for a health specialist to be available (13). A serious challenge in the use of apps, however, is that there is a general lack of evidence-based solutions, and that health apps often wrongly claim to be evidence founded (14–16). Another challenge is that the effectiveness of the majority of health apps are fairly unknown and their ability to make actual behavioral change is often poorly reported (17,18). The high availability of health apps on a unregulated market poses a major concern since it may have a major influence on rehabilitation success or at worst cause harm (14).

Injurymap© is an exercise app designed for treating different musculoskeletal problems including LAS. The Injurymap© exercise program has been developed by health care professionals and has the potential to provide an easy-accessible management of rehabilitation. However, the app has currently not been tested in a clinical study. Before undertaking a large-scale study, we wish to pilot test the app to assess the use and preliminary effect of the app-based exercise program. We are particularly interested in the proportion of patients who become active app users (and for how long) when provided with an option of a free app-based rehabilitation program in an acute care clinical setting.

### Purpose Statement

The purpose of this pilot study was to investigate the use and preliminary effect of an app-based exercise program in patients with LAS seen in the Emergency Department at a public hospital. Consistent with the mixed-method study design outlined below, both quantitative and qualitative outcomes were collected to fully explore factors related to uptake and adherence to the exercise program.

### Study Design

The study used an explanatory sequential mixed method cohort study design (QUAN→qual) (19). The study was conducted in subsequent phases, with the development of the quantitative outcomes leading the following development of qualitative outcomes, and the quantitative app-use data guiding a purposive sampling for semi-structured interviews (20). Quantitative and qualitative outcomes were analyzed separately and integration was done in the interpretation process by triangulation (sections with integration: Methods, Results, and Discussion) (21). The study process is outlined in figure 1. We consider the study exploratory and, hence, it was designed with a flat outcome structure using multiple evenly valued outcome measures.

**Figure 1:**
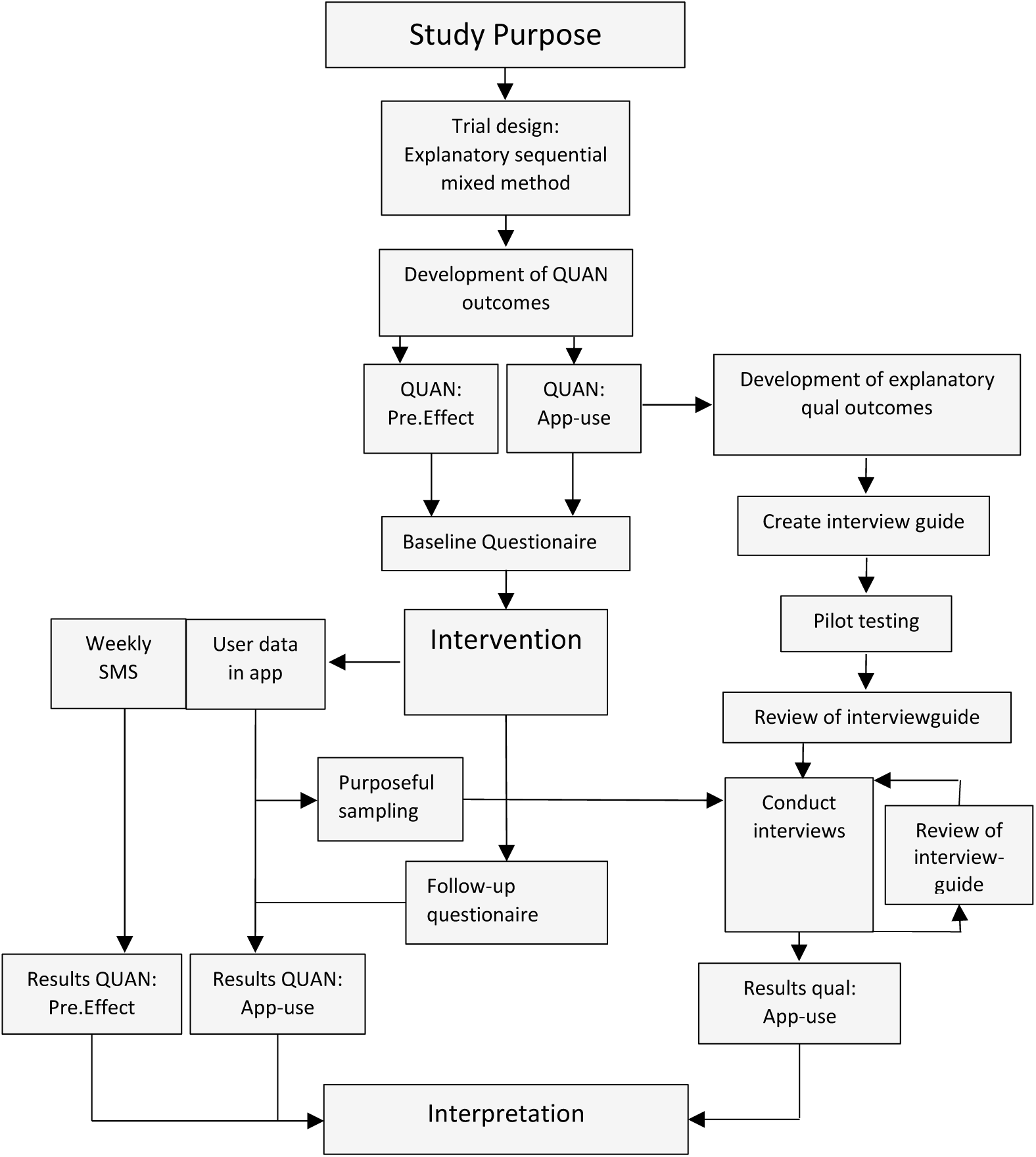
Mixed-method study process.

Outcomes related to “app-use” were quantitative data collected directly from the app during the exercise period and after by a follow-up questionnaire. Outcomes meant to provide explanatory insight of the app-use data were qualitative and collected by semi-structured interviews after the exercise period. Outcomes related to “preliminary effect” were quantitative clinical recovery-data collected using weekly TEXT-MESSAGES for four months after the initial injury.

The SPIRIT checklist (22) and PREPARE Trial guide (23) were used to develop the study protocol. The Danish National Committee on Health Ethics approved this study (ID: 17041467). All personal data were handled according to the Danish act concerning processing of personal data. Prior to the study, Injurymap had been approved for handling personal data by the Danish Data Protection Agency (file no. 2016-42-3535). The study followed the principles of the Helsinki declaration and it was pre-registered at ClinicalTrials.gov (https://clinicaltrials.gov/ct2/show/NCT03550274). We report the study using the Good Reporting of A Mixed Methods Study (GRAMMS) checklist (24) (Supplementary A).

## Method

### Study setting

The study was performed at a Danish public hospital where patients are covered by the Danish healthcare system. At the hospital, the current practice for non-surgical management of LAS patients is RICE (Rest, Ice, Compression, Elevation), mobility exercises and recommendations of slowly returning to activity. The current practice does not involve any on-site systematic instruction in evidence-based rehabilitation programs or referral to such elsewhere. Therefore, we wanted to explore the use of a simple app-based solution in this setting. Informed written consent from the participants were registered before participants started the app-based exercise program. Participants could withdraw from the study at any time, without any consequences.

### Participants

Participants were recruited from the Emergency Department at Copenhagen University Hospital, Hvidovre. They were asked to participate in a home-based rehabilitation delivered by an app (Injurymap©) available on any smart device. The inclusion criteria was; patients with an acute lateral ankle sprain (< 48 hours from injury) diagnosed by a relevant health care professional at the hospital ED. Gradings of ankle sprain severity was not performed, since this is not standard procedure in the ED. The exclusion criteria were; concurrent fracture of the leg or foot (Ottawa rules and/or x-ray), previous surgery in the ankle or surgery as a consequence of the current ankle sprain, serious illness (terminal patient, rheumatoid arthritis, fibromyalgia etc.), not owning a smart device (phone or tablet) or unable to understand and read Danish.

### Procedures

Health care professionals associated with the ED and responsible for ankle examinations recruited participants. When a health care professional identified a patient with ankle sprain, they delivered a recruitment bag containing several rubber bands of different thickness and a description of the free app-based exercise opportunity in this study. This approach was chosen to resemble a delivery method applicable in clinical practice. Besides the written information, the health care professionals were encouraged to recommend the exercise program to the patients. If a patient was willing to participate in the project, they contacted the research assistant (JB) by the contact information in the written material. When contact was established, and participants were deemed eligible, they received a voucher for free access to the app program, informed consent, and a baseline questionnaire.

The project was implemented at the ED by the primary investigator (JB). Health care professionals in the ED were informed about the study at staff meetings, by the weekly newsletter, and by the primary investigator who participated in the daily routines prior to recruitment for the project. Two large boxes containing the recruitment bags were placed strategically in the ED office and the primary examination room. The boxes had a large picture of an ankle sprain at the front and a text asking to give patients an exercise opportunity. This recruitment procedure was chosen to reflect a normal clinical care setting and to encompass both the health care professionals’ willingness to promote the app solution to patients with LAS, as well as participants’ willingness to accept the offer. For the same reason, we tried to inform about the study as being a rehabilitation exercise opportunity more than a research study.

### Intervention

InjuryMap© offers exercise programs for LAS and other musculoskeletal conditions. The app requires user-registrations and a monthly paid subscription fee to access the exercise program. In this study, the app company provided free access for participants to the LAS exercise program. Examples from the app content can be seen in figure 2.

**Figure 2:**
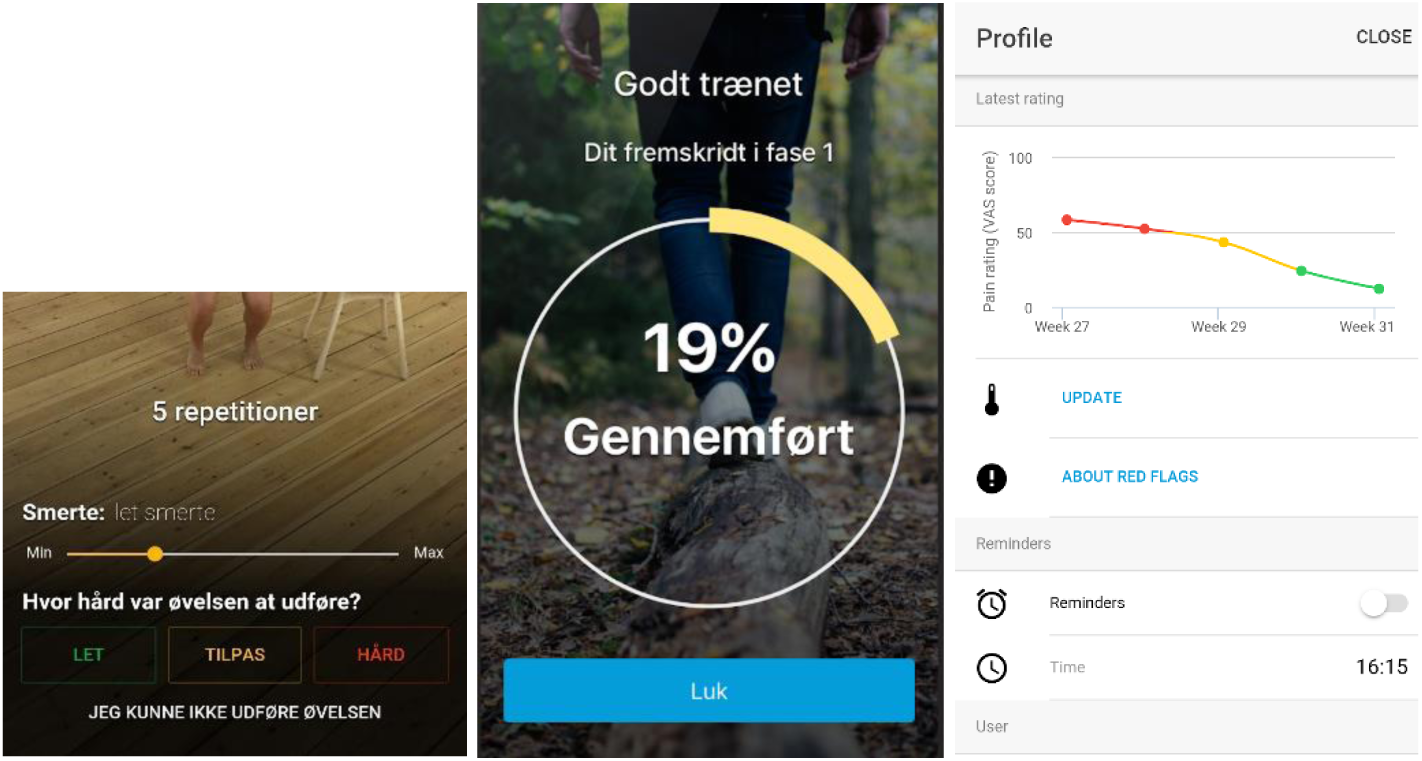
**Example of app content. medRxiv policy does not allow for publication of images that may be identifiable to the individual, their friends, family, or neighbors. Pictures are available from the corresponding author and from the peer-reviewed paper, once published.**

The exercise program was available on any mobile device and/or tablet using Android or iOS operating systems. Participants could perform the exercises at any preferred location and were not restricted from seeking additional care. After four months of training or if a participant was inactive in the app for more than two consecutive weeks, they were considered to have stopped the exercise intervention.

The exercise program for LAS consisted of three phases with increasing difficulty. Each phase consisted of four categories of exercise. The categories were 1) mobility, 2) stability/balance, 3) strength and 4) stretching. The app could adjust the difficulty for each individual exercise depending on user-feedback. A comprehensive description of the exercises can be found in Supplementary B.

The exercise program was set up so that several exercises were completed after each other to successfully complete a training session. Each exercise was accompanied by a video with an explanatory voice-over and the number of required repetitions written on the display. After completing an exercise, the participants registered pain level and difficulty of performing the exercise. If the participants registered no or low pain and low exercise difficulty, the app chose a progression of the exercise for the next exercise session. The app recommended participants to complete exercise sessions three times a week after app registration. If the participants followed the recommended three sessions a week, they would be able to reach the highest difficulty level in two to four months depending on their pain and difficulty answers. There were no limitations in how many exercise sessions could be performed per week or a maximum number of weeks they could exercise except for the strengthening exercises which could only be performed once per day. Participants were able to activate a reminder function so that the app would remind them to exercise on a daily timepoint of their choosing. The spoken and written language in the application was Danish.

In the exercise program, circular rubber bands were used in several exercises. The rubber bands are common and cheap products available in most sports stores. In this study, the recruitment bag contained a selection of rubber bands with varying resistance.

### Outcomes

In this study, the outcome data were divided into two categories reflecting the study objective. The first category (“App-use”) consists of quantitative data on uptake, retention, and adherence. Furthermore, it contained user-experience which comprised of quantitative data on satisfaction with the app and qualitative data on the factors that influenced the app use. The second category (“Preliminary effect”) consists of quantitative data on clinical recovery and recurrent injuries. A baseline questionnaire was completed after enrollment to gather descriptive data.

#### App-use

The overall rationale for the app-use outcomes below was to investigate how many people exposed to the app that started using it; how much they used it; when they stopped using it; and how the user experience was. As a part of the app evaluation, we assessed uptake of the app-based exercise program. For app uptake, we calculated the following: number of people diagnosed with an ankle sprain at the ED in the study period; number of people who received a recruitment bag; number of people willing to participate (contacted by the principal investigator); number of people who became active users (defined as having downloaded the app and initiated the exercise program). By counting how many recruitment-bags the health care professionals delivered, the number of people with ankle sprain who had been informed about the exercise opportunity could be estimated. From the Danish National Patient Register it was possible to obtain the number of ankle sprain diagnosed at the ED. For retention, following were calculated: Number of people completing baseline and follow-up questionnaire; and number of text-messages responded through the study period.

The app had mandatory user-registration so all exercise activity in the app was registered at the individual participant level. From these data, we calculated the following adherence outcomes: Number of exercise sessions completed per participant; and completed exercise sessions per week. If a participant didn’t commence the exercise program within two weeks, when active users was inactive for two consecutive weeks, or if they were active in the app for four months from their initial injury, they were considered finalized and received the follow-up questionnaire containing user-experience with the app-based exercise solution. Satisfaction on a five-point Likert scale were assessed for active users for the following items: the difficulty and the progression of the exercise program (Difficulty), the content of the exercise program (Content), the results from the exercise program (Results), and the usability of the app (User-friendly). Furthermore, all participants were asked if they would recommend the app to others (yes/no) and how much they would be willing to pay for the app (DKK).

After ending the exercise intervention, a group of participants were contacted for semi-structured interviews. The interviews focused on understanding and explaining motivational factors or barriers that may influence the use of the app “Injurymap©”. The study used a purposeful sampling for the interviews as recommended for explorative mixed method studies (20). The sampling of participants was based on different number of completed exercise sessions, different age groups, both men and women. We did this to capture get a broad perspective on the motivational factors or barriers from both those with many completed exercise sessions and those who dropped out early. Based on the sampling criteria, a pragmatic number of ten participants were selected (both men and women at different age groups).

The interviews were performed by phone by the principal investigator (JB). Participants for the semi-structured interviews were contacted by mail and phone at the same time as the follow-up questionnaire. An interview guide was developed by the principal investigator (JB) with supervision from a senior researcher experienced in qualitative research (JWK). The guide was pilot tested on a person with experience in the exercise app, but otherwise not involved in the study. Recordings from the pilot testing were examined by two researchers (JWK and JB) to improve the interview technique and evaluate coherence of the questions in the interview guide. After the first interview, the recordings were examined again by the senior researcher (JWK) the recordings were compared to the purpose statement to ensure it was adequately covered in the interviews. Changes in the interview guide from the first interview consisted primarily of merging separate themes, based on how the participants associated and described their experiences. We also added questions about the experience at the ED and its impact on app-use since this factor was mentioned as an influential factor.

Interviews were recorded and transcribed verbatim. The data were analyzed using a thematic approach as described by Castleberry (25). The data were coded, and recurring phrases or words were grouped into basic themes by the principal investigator (JB). Themes and codes were compared to the transcribed interview by two researchers (JB and JWK) to ensure that coding was performed with the same consistency and true to the original statements. This was performed in several processes until agreement was achieved. In this process, basic themes were clustered into global themes. The initial interpretation was performed by the principal investigator (JB) and reviewed by a researcher (JWK). Finally, the qualitative results were discussed with the whole research group.

#### Preliminary effect

The overall rationale for the preliminary effect-outcomes outlined below was to investigate if exercise adherence, was related to clinical recovery. Clinical recovery was evaluated by self-reported evaluation of symptoms using a weekly string of text-messages for four months after their initial injury. The following Clinical Recovery items were collected by text-messages: Not able to fully participate in work/study because of the ankle sprain (days); Return to sport (RTS), defined as not able to fully participate in sport because of the ankle sprain for participants who registered as being “sports active” in the baseline questionnaire (weeks); Recurrent lateral ankle sprains in the same ankle (number); Subjective feeling of ankle stability (0-10 points). From the follow-up questionnaire, the clinical recovery item: Subjective feeling of recovery (yes/no) was also collected.

A recurrent sprain was defined as an inversion episode on the same ankle as assessed in the ED. Recurrent sprains were divided into two groups; 1) Recurrent sprain with time-loss, defined as being unable to continue current activity and/or unable to participate in work/sports activities the next day because of the ankle; 2) Recurrent sprain with no time loss was defined as able to continue with current activities and able to participate in sports/work activities the next day.

As the exercise program in the app is built with similar component as other evidence-based exercise programs (8), we expected no harms from the intervention. Nonetheless, participants received a text-message in the weekly string of text-messages where they could register any discomforts or injuries related to performing the exercise program.

### Materiel and outcome assessment

The baseline and the follow-up questionnaires were collected without assessor involvement through RedCap® (Research Electronic Data Capture) - a browser-based software developed by Vanderbilt University. Participants received mails containing a link to their personal online questionnaire. The research team had access to app use data through a log-in to the Injurymap© online database. The text-messages were collected using SMS-track© - an online system used to send and receive standardized text-messages. Each week the SMS-track system sent the first of six questions to the participants and waited for an answer before the next question was sent. If a participant reported an ankle sprain, she or he received a phone call from the principal investigator (JB) to clarify the type of sprain.

The principal investigator (JB) performed the outcome assessment, follow-up assessment, data extraction and data analysis. Participants active in the app were contacted as little as possible to minimize any potential influence on their exercise behavior.

### Statistical analysis

Baseline characteristics are summarized using suitable descriptive statistics. Normal distribution was assessed using the Shapiro-Wilk test and Q-Q plots. For the app-use outcomes, recruitment rates, retention rates are presented in suitable descriptive tables. Adherence is summarized in total sessions per participant and exercise sessions per week during the intervention period. Registered Harms were addressed individually. Quantitative user-experience are summarized using descriptive statistics.

We planned to determine the preliminary effect of the app by examining the relationship between the exercise dose (adherence) and clinical recovery outcomes using linear or logistic regression models, depending on type of outcome. The models would include the clinical recovery outcome as the dependent variable and exercise dose as independent variable. However, as the weekly text-messages that contained questions on clinical recovery were answered more frequently by participants who were very active in the app, we chose not to conduct the analyses of how exercise adherence related the clinical-outcomes as they would be biased. Instead, we report the clinical recovery data using descriptive statistics.

### Sample size

Approaches to sample size justification for studies that investigate preliminary effectiveness of interventions, such as pilot and feasibility trials, vary. One rule of thumb-approach is 12 per group for a pilot RCT (26). We used a pragmatic sample size 60 participants for this study. It was based on the rationale that 30 out of 60 would download the app, start using the app, and start using the exercise program, based on previous experience with the app on low back pain patients. We figured a sample size of 30 app users would equate to two groups of 12 participants each, if pooled (26), plus 6 to account for some attrition.

## Results

60 participants were recruited during the period from July 3, 2018 to April 3, 2019. 43 of these stated that they participated in weekly sports activities (Sport active group). Accounting for half of the sprains, sports were the most frequently reported cause of injury. One third (n=20) reported a previous ankle sprain in the same ankle. The sprains were equally divided between left and right site sprains. Baseline characteristics are presented in table 1.

**Table 1:** Baseline characteristics.

| **Item** | **Mean(SD)** | **N** |
| --- | --- | --- |
| **Age (yr)** | In their 30s | 60* |
| **Weight (kg)** | 76,60(19.85) | 56 |
| **Height (cm)** | 174.04(10.75) | 56 |
|  | **n(%)** | **N** |
| **Women** | 36(64.3%) | 56 |
| **Injury site (Right)** | 29(51.8%) | 56 |
| **Previous (same) ankle sprain (yes)** | 20(35.7%) | 56 |
| **Sports active, (yes)** | 43(78.2%) | 55 |
| **Education level:** |  | 56 |
| *Primary education* | 11(19.6%) |  |
| *Upper secondary education* | 10(17.9%) |  |
| *Vocational Education* | 4(7.1%) |  |
| *Short higher education* | 1(1.8%) |  |
| *Bachelors program* | 23(41.1%) |  |
| *Master program or higher* | 6(10.7%) |  |
| *Other* | 1(1.8%) |  |
| **Physical demands on work:** |  | 55 |
| *Mostly sitting* | 22(40%) |  |
| *Equal sitting and walking* | 19(34.5%) |  |
| *Mostly Walking* | 14(25.5%) |  |
| **Activity when injured:** |  | 56 |
| *Work* | 8(14.3%) |  |
| *Sports* | 26(46.4%) |  |
| *Leisure* | 22(39.3%) |  |
| *Other* | 0 (0%) |  |
| **One or more element of the RICE-principle (yes)** | 52(92.9%) | 56 |

*\*Data on age were collected via the written informed consent. All other data were collected via the baseline questionnaire. Wording chosen to comply with medRxiv's policy regarding limiting the number of indirect identifiers.*

Characteristics of the 10 participants interviewed can be seen in table 2. They had completed between 1 and 26 exercise sessions. 19 people were contacted to achieve 10 interviews. One person refused to participate in the interview due to lack of time. The remaining eight did not respond to the request.

**Table 2:** Characteristics of the interviewed participants.

| ID | 2 | 4 | 5 | 7 | 8 | 9 | 12 | 13 | 17 | 23 |
| --- | --- | --- | --- | --- | --- | --- | --- | --- | --- | --- |
| M/W | W | W | M | M | M | W | M | M | M | W |
| Age (yr) | * | * | * | * | * | * | * | * | * | * |
| Weight (Kg) | 78 | 90 | 75 | 112 | 95 | 65 | 91 | 80 | 84 | 56 |
| Height (cm) | 164 | 165 | 166 | 190 | 181 | 168 | 184 | 192 | 186 | 164 |
| Education<sup>a</sup> | PostG | PostG | Skill | Grad | Grad | Grad | Grad | Grad | <High | <High |
| Employment | Out | Job | Job | Job | job | Job | Out | Job | job | Stud< |
| No sessions completed | 26 | 21 | 4 | 16 | 8 | 3 | 5 | 1 | 7 | 3 |

<sup>a</sup>Education: < High School (<high), High School (High), Skilled (Skill), Graduate (Grad), Post Graduate (PostG)
<sup>b</sup>Employment: Employees or self-employed (job), Unemployed (No), Not in the labor force (out), Student in high school or lower education level (Stud<), graduate student or higher education level (Stud>). \* Removed to comply with medRxiv's policy regarding limiting the number of indirect identifiers.

From the coding process (25), three themes were deducted (see table 3). Each theme was divided by several sub-themes. Theme I-II are characterized by a substantial amount of data and a clear link to the purpose of the interviews. Theme III is also characterized by a substantial amount of data and includes factors that were not directly linked to the app experience but with a potential impact on the use of the app. Quantitative adherence data and qualitative data are presented through joint display table as recommended by Guetterman et al. (27).

**Table 3:** Themes and subthemes.

| Themes | Subtheme |
| --- | --- |
| **I: Motivational factors** | *Usability*<br>*The app's exercise level and ability to adapt*<br>*Ankle symptoms and expectations*<br>*Influence of work and leisure time* |
| **II: Technology assisted exercise behavior** | *Process statistics*<br>*Reminder function*<br>*Exercise Comprehension*<br>*Views on the visual expression* |
| **III: Factors of importance for start-up** | *Diagnostic and prognostic expectations*<br>*Treatment and preventive expectations*<br>*Provider integrity*<br>*The user as independent searcher* |

### App-use

#### Uptake

According to the Danish National Patient Register (28), a total of 1110 people were diagnosed with an ankle sprain in the ED during the study period (see table 4). It should be noted that this does not only include isolated sprains or lateral sprains, which is why the number of people is considered an absolute maximum of potentially eligible participants. 485 people received the recruitment bags containing rubber bands and study information. Of these, 60 contacted the principal investigator and were included. 48 became active users of the app, according to our definition.

**Table 4:** Uptake of the app intervention.

| Project stage | No of people | Percentage of total diagnosed | Percentage of those who received info |
| --- | --- | --- | --- |
| Diagnosed with ankle sprain in the ED | 1110 | 100% | - |
| Received study information | 485 | 44.7% | 100% |
| Enrolled in the study | 60 | 5.4% | 12.4% |
| Active users | 48 | 4.3% | 9.9% |

#### Retention

Of the 60 people enrolled, 54 (92%) answered the baseline questionnaire (one participant had a partial completion) and 46 (77%) answered the follow-up questionnaire. Two people dropped out, one because he did not become an active app user and, hence, did not want to answer the SMS-string, one due to pregnancy.

For the 60 people in the 17 weeks follow-up period, a total of 6120 SMSs could potentially have been answered. A total of 4387 answers were received (72%), with the highest frequency in week 1 (85.0%) and the lowest in week 13 (61.9%) (see figure 3).

**Figure 3:**
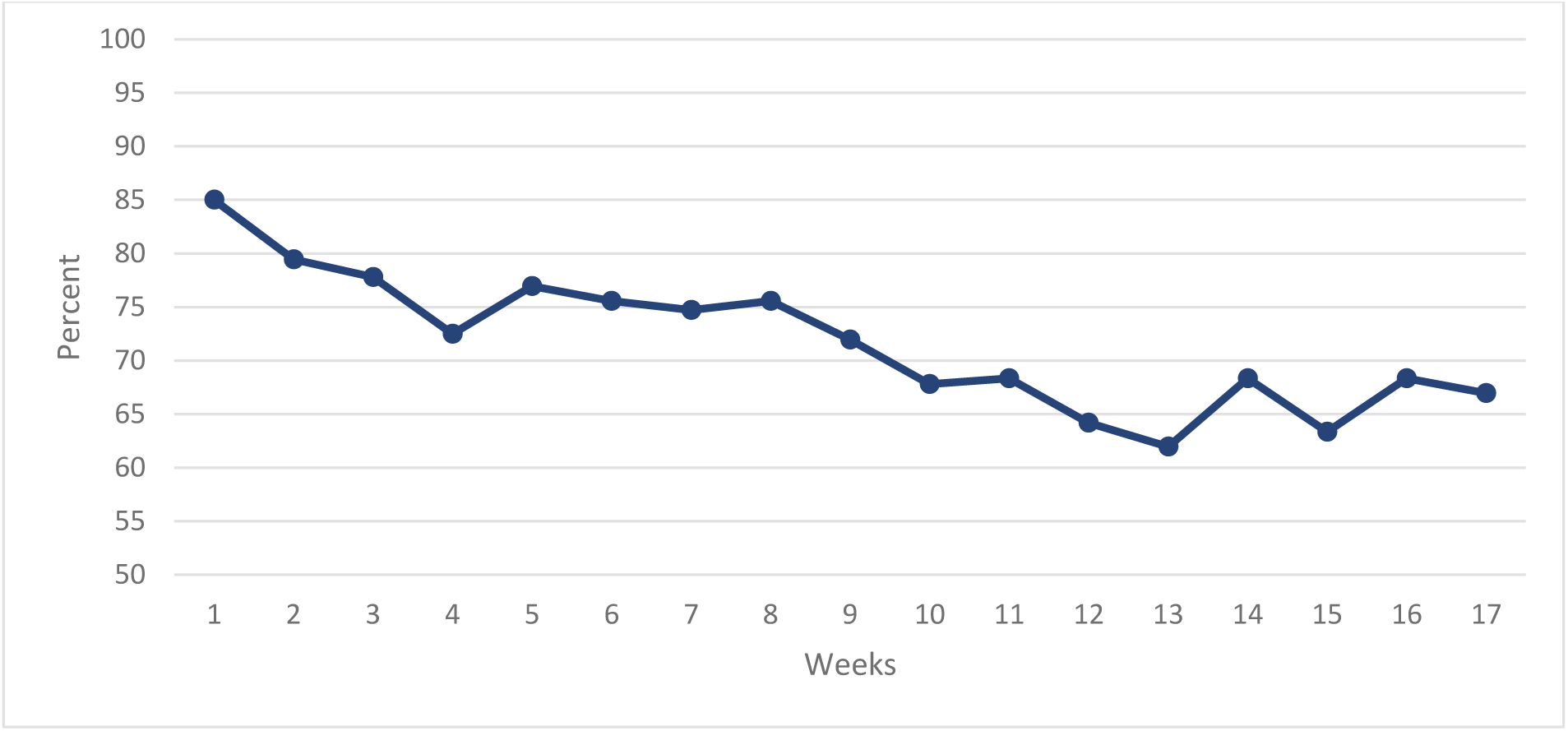
Weekly SMS response rate.

#### Exercise adherence

48 participants became active users because they completed a minimum of 1 exercise session (see figure 4). The median number of completed exercise sessions in the four months period was 5.5 and ranged from 0 to 68 completed sessions (see Supplementary C). An exploratory analysis of the adherence by education level, age, work, and sports active can be found in Supplementary D.

**Figure 4:**
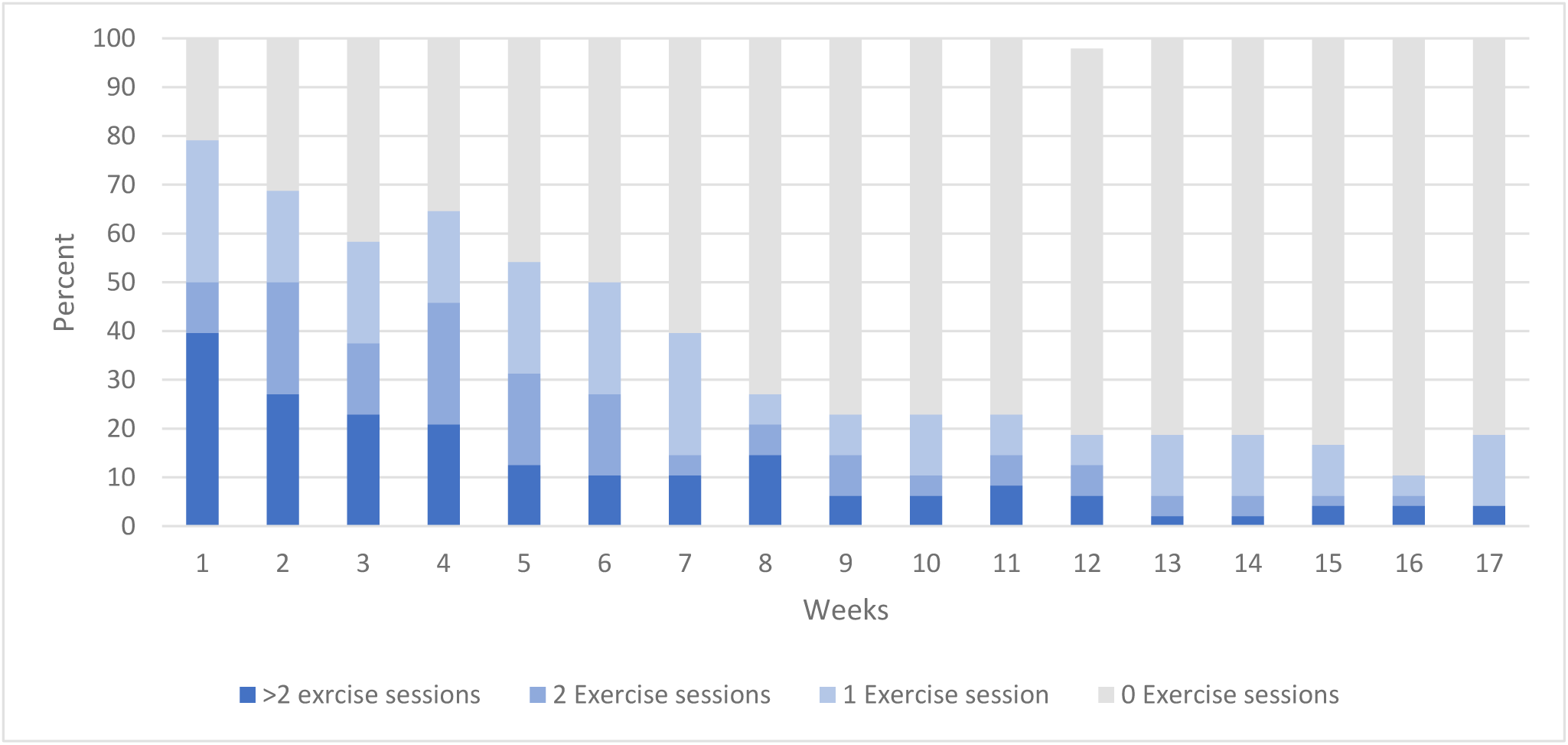
Weekly distribution of completed exercise sessions.

The interviews sought to gain an explanatory insight regarding factors that may have influenced uptake and adherence to the exercise program. Theme I “Motivational factors” describes factors directly linked to adherence by the participants. Theme II “Technology assisted exercise behavior” describes how participants viewed technological features that may have influenced adherence but was not directly connected by the participants. Theme III “Factors of importance for start-up” describes factors that may have influenced participants to become active users. See table 3 for subthemes related to the themes and table 5 for a joint display of the quantitative and qualitative findings.

**Table 5:**
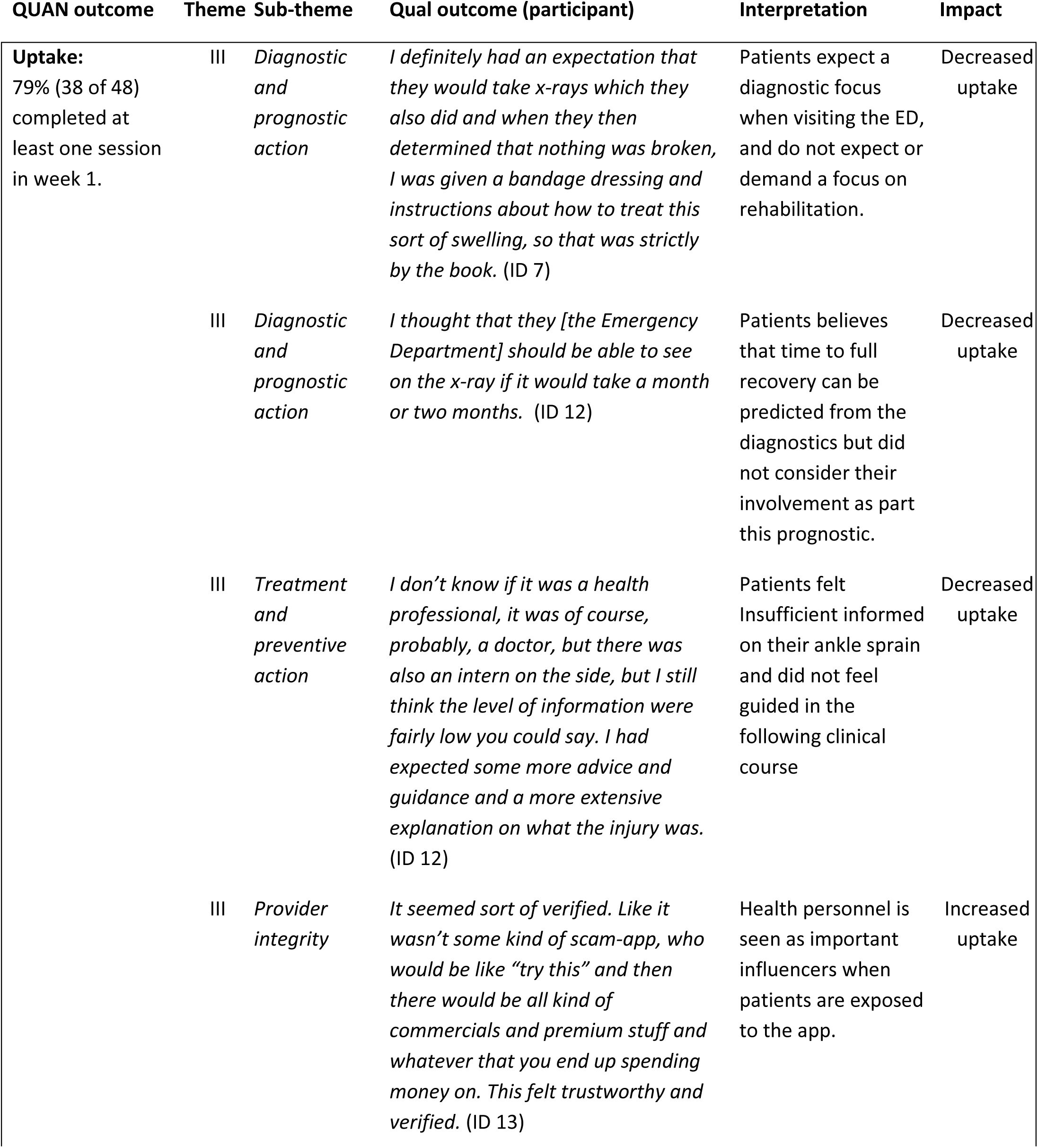
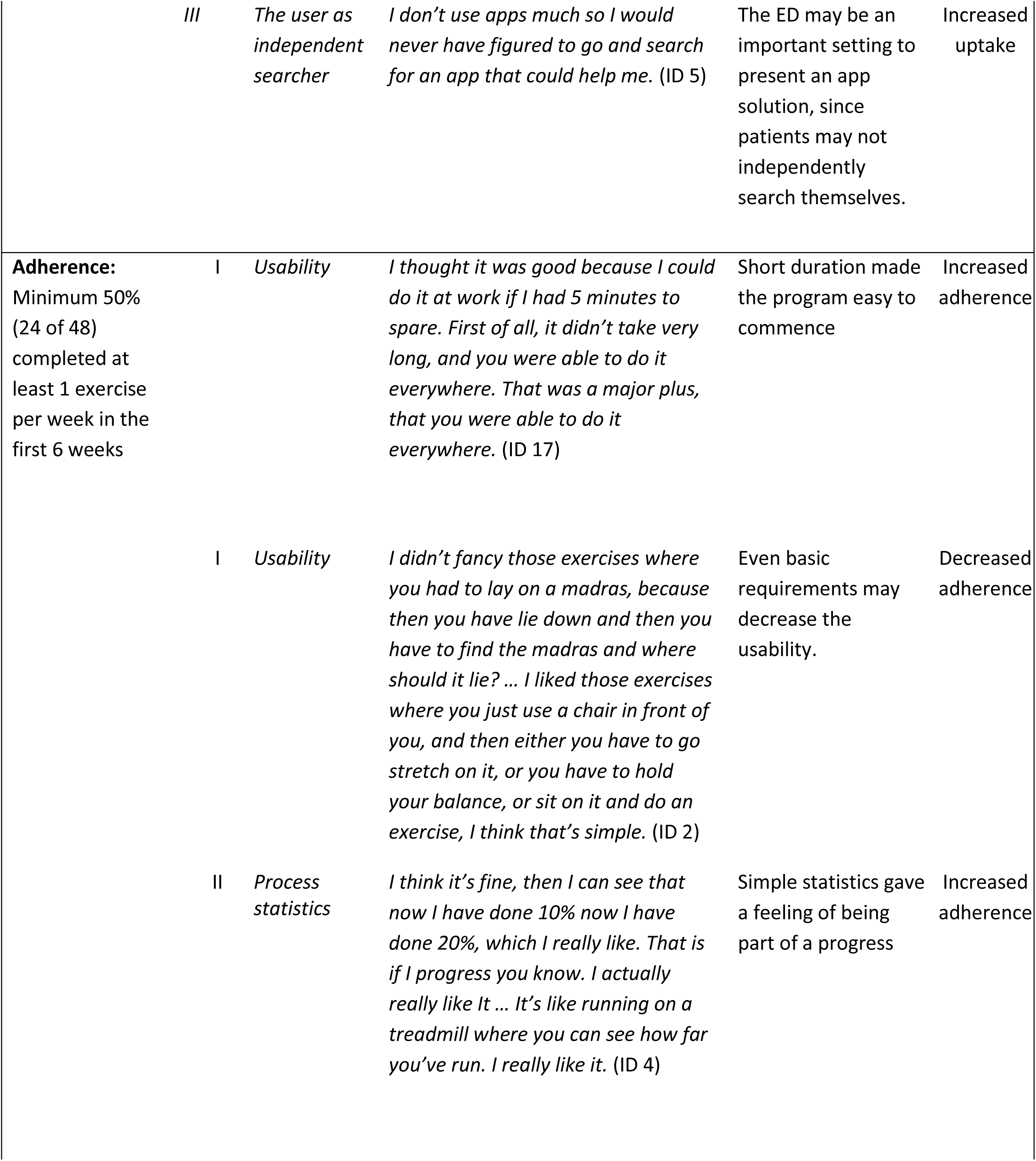
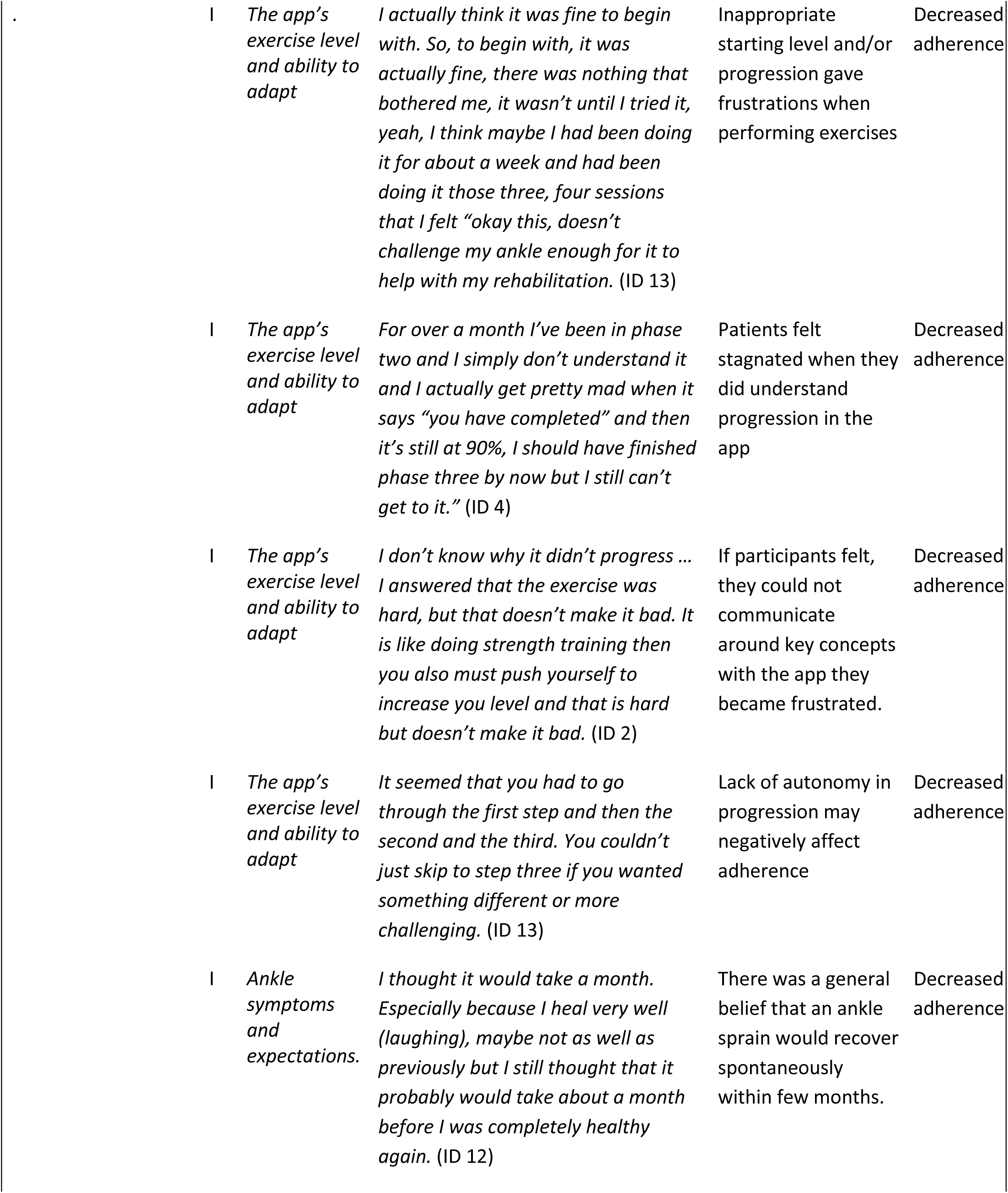
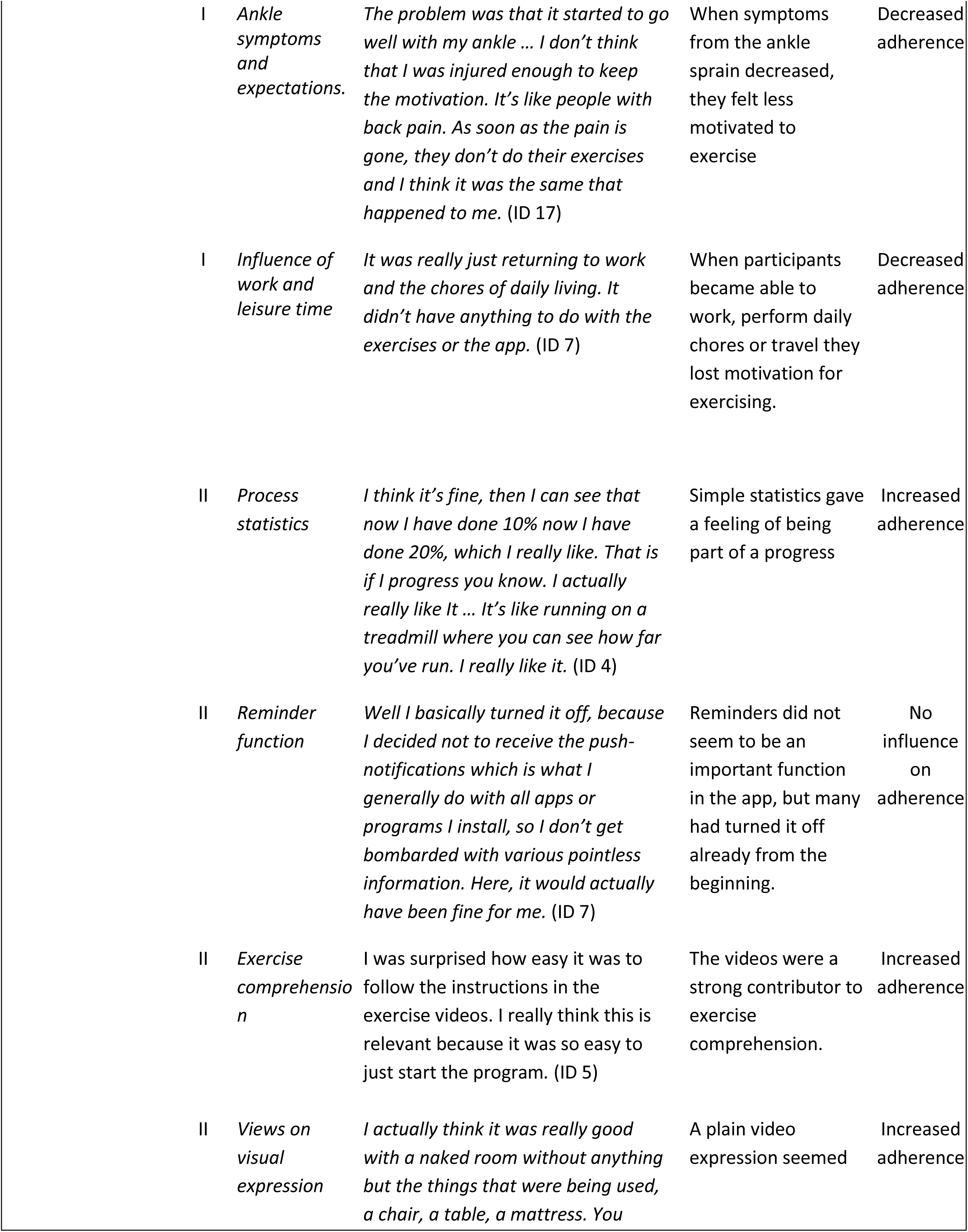
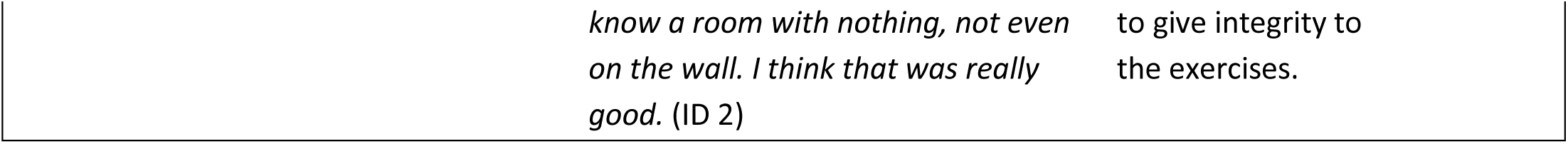
Joint display of quantitative adherence data and qualitative explanatory findings.

### Satisfaction

When asked at follow-up, 95.7% of the participants would recommend the app to other people with an ankle sprain. 71% were willing to pay for the exercise program with an average cost of 46 DKK equivalent to 6.16 EUR. Satisfaction scores can be seen in figure 5.

**Figure 5:**
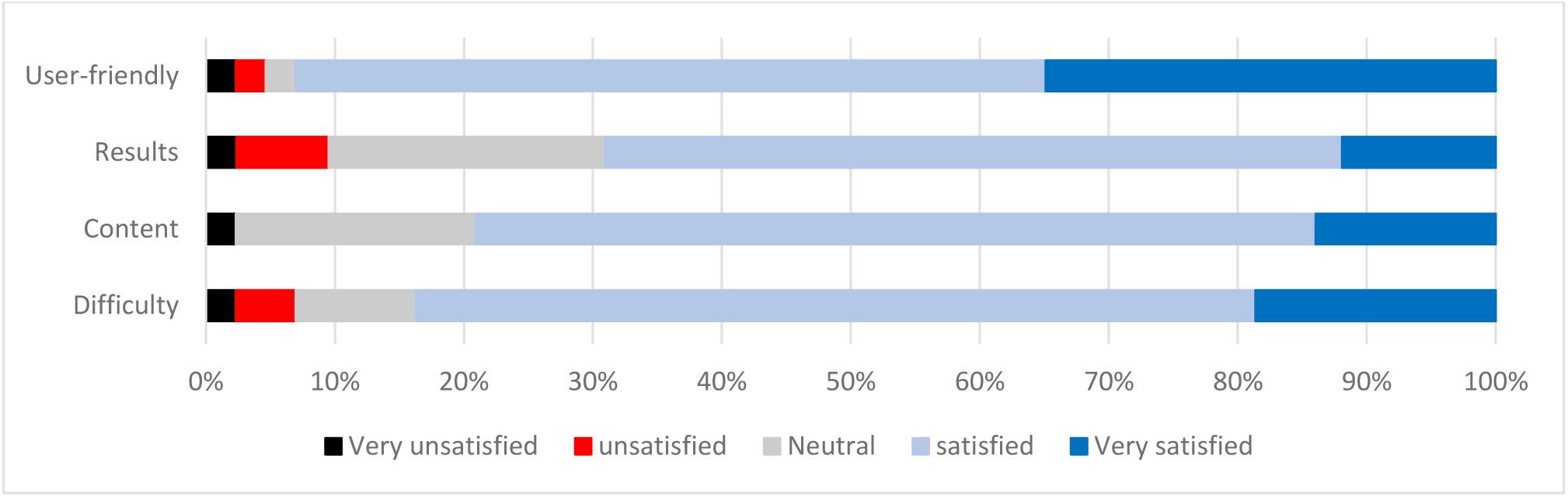
Satisfaction scores for different app use items.

### Preliminary effect

A total of 36 recurrent sprains were reported in the follow-up period. Of these, 32 were time-loss injuries and 4 were not. 20 participants had minimum 1 recurrent sprain and 11 of the 20 had 2 or 3 recurrent sprains. No participants had more than 3 recurrent sprains in the period. At follow-up, 39.1% (18 people) felt that the ankle was able to perform at the pre-injury level. The absent from work/study ranged from 0 to 100 days, the median being 1 (IQR=6) day. In week 1 the average ankle stability was 4.5 (SD=2.5) on the 0-10 scale and increased to 8.7 (SD=1.5) in week 17. Weekly changes can be seen in table 6. The sports active group (n=43) had in average 9 weeks (SD=4.9) where they could not participate in sports activities without restrictions from their sprained ankle. After 17 weeks, 30.2% still reported that they were restricted by their ankle in sports activities.

**Table 6:** Weekly ankle stability scores.

| Week | 1 | 2 | 3 | 4 | 5 | 6 | 7 | 8 | 9 | 10 | 11 | 12 | 13 | 14 | 15 | 16 | 17 |
| --- | --- | --- | --- | --- | --- | --- | --- | --- | --- | --- | --- | --- | --- | --- | --- | --- | --- |
| Ankle stability | 4.5 | 5.7 | 6.5 | 6.8 | 6.9 | 7.4 | 7.5 | 7.8 | 7.9 | 8.2 | 7.9 | 8.1 | 8.1 | 8.6 | 8.6 | 8.6 | 8.7 |

*The mean scores for subjective ankle stability (0-10 points) for each week. 0 = Very unstable, 10 = Completely stable.*

### Harms

No harms were registered.

## Discussion

The present study investigated the use of an app-based, rehabilitation exercise program for lateral ankle sprains by collecting data on use of the app “Injurymap”.

From the 1110 patients who were diagnosed with an ankle sprain at the ED during the study period, 45% received the information about the free app-based exercise program. Of those who received the information, 10% became active users. In this study, we were able to determine exactly how many received study information and how many became active users. To our knowledge, this is the first study to provide a precise estimate of how likely people are to use a rehabilitation app when presented with the opportunity in a clinical setting. This, in turn, allows for evaluation of app uptake and if changes in recruiting method or the app design affect use. We consider such data important in understanding the expanding and unregulated field of health apps (14). Since this is the first study to evaluate the delivery of an exercise app for ankle sprains in an ED setting, it is difficult to compare the uptake data to many other studies. Vriend et al. (29) did evaluate the uptake of an exercise app for people with previous ankle sprains when advertised in national media and on sports facilities. They estimated that the app was downloaded by less than 2.6% of their targeted population despite intensive marketing, and that only 62% of those who downloaded the app became active users. Though the authors were not able to determine the percentage of the targeted population that became aware of the app’s existence, they concluded that a “marketing” type of strategy may not be the optimal method of implementing an evidence-based app. Compared to the estimated 2.6% referenced above, the 10% active users in our study was better, however, it is difficult to consider 10% uptake a success. The higher uptake-level in our study indicates that the direct delivery of the app from a health care professional in the ED can encourage more people to download the exercise program. This was further supported by our qualitative data showing that the app, when given by a health care professional, seemed trustworthy and this had influenced participants to download the app; a finding that aligns with a previous study where people stated they were more likely to use an app if it was endorsed by a health care professional (30). It was, however, beyond the scope of this study to investigate how the health care professionals delivered the app information and their beliefs towards it, and whether this affects uptake to a high degree. Interestingly, the interviews also revealed that several participants felt insufficiently informed about their injury when leaving the ED. This might indicate that more adequate information from health care professionals about the consequences of LAS and recommended exercise rehabilitation could prompt more patients to use the app and health care professionals in the ED could have a great opportunity to influence people’s behavior by providing such information in a clinical setting.

The active users completed a median of 5.5 exercise sessions. Most participants became active in the first week, but 20% started after the first week. In general, adherence declined through the study period and after 2 weeks less than half of the users performed 2 or more weekly sessions. After 9 weeks around 20% continued to use the app through the 8 months study period. Because this study was exploratory, we did not pre-specify a threshold for acceptable adherence. This is important because studies have found that a home-based exercise program with 24 exercise sessions could reduce the risk of recurrent sprains by 35% (12,31). If we consider 24 exercise sessions to be an acceptable adherence threshold (not considering time per session or time intervals between completed sessions) only 15% of the participants in the present study were adherent.

Despite the low adherence, the participants reported high satisfaction with the app. Almost all participants would recommend the app to others, which is consistent with data from Vriend et al. (29). Their app was also given high appraisal by its users even though they only completed, on average, 3.3 exercise sessions out of the recommended 24 in the app. One would think that they stopped exercising because they no longer felt restricted by their ankle sprain. However, when asked about this in the present study (if they felt recovered when they stopped using the app) the majority responded “No”. Participants stated in the interviews that they had lost motivation when the symptoms declined to a level where they were able to manage daily tasks. From the clinical recovery data, we can see that crutches are predominantly used in the first two weeks. So, even though 80% of the participants after two weeks still suffered symptoms, it is likely that they could have started to work and participate in leisure activities and thus feel it less necessary to continue using the app. This may have contributed to why they stopped exercising with the app.

The interviews revealed that participants in general expected a complete spontaneous recovery from the ankle sprain regardless of their actions. That people with ankle sprain believe the injury to be innocuous is anecdotally supported by several studies (1,4,10,32–34) but to our knowledge, this is the first study that has interviewed people on this perception. The perception that an ankle sprain is an innocuous injury may also be reflected in the reason for seeking medical attention at the ED, as participants primarily went to the ED because they were worried that the ankle had a fracture, not because they were worried about the consequences of a sprained – not fractured – ankle. How the perception of LAS as being an innocuous injury influences adherence to an app and an exercise program is unknown. This knowledge could potentially provide a means to increase the effectiveness of such an exercise intervention.

The interviews revealed that several participants experienced the starting level to be either too difficult or too easy. They found it discouraging if the app did not match their expectations regarding exercise difficulty within a few completed sessions. The app was designed with a fixed starting level and a hierarchical development of exercises. The advantage of this design is that the progression can be matched to exercise guidelines and minimize the risk that users are presented with exercises that may cause them harm. The disadvantage is that some participants may need to complete several sessions before they reach a desired exercise difficulty, which may negatively impact adherence.

The app did provide the participants with an exercise solution they found easy to access and the short programs were perceived as easy to include in the daily routines. Both time restrictions and access to exercise opportunities have been found to be major barriers for physical activity among patients with musculoskeletal disorders (35). It is interesting that despite the app-based exercise program resolved these two major barriers and it was highly appraised, it was not enough to substantially motivate our participants to exercise. Whether the low adherence was primarily due to a general opinion that ankle sprains are an innocuous condition, and/or the app-based exercise program – especially the starting level – is currently unknown. Further research is needed to evaluate how different recruitment methods, program designs and conditions affect adherence for app-based exercise interventions

## Future app optimizing

The app-based exercise program enables users to perform exercises wherever and whenever. However, data from the interviews point out that usability of an exercise app may be more than just being ever-present in your pocket. Time to complete sessions, exercise materials, the need for changing into gym clothes or just getting down on the floor are all factors that may influence and limit the usability. Some demands can be necessary for the exercise program to ensure correct exercise form; however, it may enhance adherence if users could customize their program to fit not only their injury but also their exercise behavior. Giving users more control of their exercise program may also be a solution for users who feel that the starting level is far from what they want it to be. The exercise app seemed to have one or two sessions to match people’s expectations before they would quit the program. Since the app can make real-time changes dependent on user feedback, an improvement could be to include initial questions on people’s functional disabilities (e.g. ability to stand or walk), so that it may guide the app to a more motivational starting difficulty. Furthermore, giving users the ability to skip difficultly levels would make the app more adaptable to both user expectations and day to day changes in symptoms.

It seems that the explanatory exercise videos are important, and they seem to make people feel secure in their exercise execution. The visual expression in the videos seems to influence the integrity of the app. Most of our participants seemed to prefer what they called the “clinical expression” used in this app with a bare room and a regular looking person, compared to more fitness-focused app with highly trained athletes and fast paced music. They wanted a person they could relate to. It is likely that different age groups, gender etc. prefer different video expressions and a personalized video could enhance motivation to perform exercises with the app as a partner.

With regards to the statistics in the app, we were surprised that participants did not consider them important for their motivation. It was surprising because many thought the process statistics presented after each completed session were a measure of their clinical recovery despite the fact it only reflected the program process. This is, however, similar to the finding from Liao et al. (36) who found that visual demonstration was perceived as the only important motivational factor among 52 app design features including reminder and statistical functions. A reason that the statistics are not found to be motivating could be that they are generic and not user tailored. A patient-specific goal setting may enhance users work towards those goals (37,38) and a scale like the Patient Specific Functional Scale (39) would be easy to include as an app feature.

## Strengths and limitations

The adherence data in this study do not rely on feedback from participants and are therefore not affected by potential recall or reporting bias. This is a major study strength. Only exercises that are registered in the app are recorded, however. One participant described that after learning several of the exercises, she had performed them without opening the app.

The app collects feedback on individual exercises, which is much more detailed than just session collecting completion, which is a commonly used proxy measures for adherence (40). From the data, it would be possible to identify possible exercises that participants experience too difficult or pain provoking at a certain stage. A limitation, however, it that the app does not monitor patients during the exercises and performance quality is not assessed.

In this study it was possible to elaborate the quantitative app-use data with qualitative data from the interviews and suggest possible explanations. This would not have been possible if only one methodological approach has been chosen. The mixed method approach in this study is described with regards to “Good

Reporting for a mixed method study” (GRAMMS) (24) to increase transparency and the author group comprised of both quantitative and qualitative experts to obtain quality of each approach which is advocated for mixed method design (24).

## Conclusion

In this study, only few of the patients seen for an ankle sprain in the ED became active app users after they received information about a free app-based rehabilitation exercise program. Those who did, liked the app very much, but few completed enough exercise sessions to realistically impact clinical recovery. The ankle sprain was generally considered an innocuous injury that would spontaneously recover even though more than half the participants did not feel fully recovered when they stopped exercising, and a third experienced a recurrent sprain. To improve the care of these patients in the ED, we suggest that health-care personnel who asses acute ankle sprains should be aware of their importance in informing patients about the risk of prolonged symptoms and recurrent sprains, so that patients may have more realistic expectations on the clinical course. Recommendations from a health care professional in the ED to use an ankle sprain exercise program seems to carry more weight than similar recommendations given elsewhere, making the ED setting interesting from an implementation point of view.

## Supporting information

ICMJE_COI_all_authors

## Data Availability

All data produced in the present study are available upon reasonable request to the authors

## References

1. Gribble PA, Bleakley CM, Caulfield BM, Docherty CL, Fourchet F, Fong DT-P, et al. Evidence review for the 2016 International Ankle Consortium consensus statement on the prevalence, impact and long-term consequences of lateral ankle sprains. Br J Sports Med. 2016;50(24):1496–505.

2. DS93: Luksation og distorsion af led og ligamenter i ankel og fod [Internet]. The Danish National Patient Registry. 2017. Available from: https://www.esundhed.dk/Registre/Landspatientsregisteret

3. McGovern RP, Martin RL. Managing ankle ligament sprains and tears: current opinion. Open Access J Sports Med. 2016;7:33–42.

4. Feger MA, Glaviano NR, Donovan L, Hart JM, Saliba SA, Park JS, et al. Current Trends in the Management of Lateral Ankle Sprain in the United States. Clin J Sport Med Off J Can Acad Sport Med. 2017;27(2):145–52.

5. Larmer PJ, McNair PJ, Smythe L, Williams M. Ankle sprains: patient perceptions of function and performance of physical tasks. A mixed methods approach. Disabil Rehabil. 2011;33(23–24):2299–304.

6. van Rijn RM, van Os AG, Bernsen RMD, Luijsterburg PA, Koes BW, Bierma-Zeinstra SMA. What is the clinical course of acute ankle sprains? A systematic literature review. Am J Med. 2008;121(4):324–331.e6.

7. Bleakley CM, O’Connor SR, Tully MA, Rocke LG, Macauley DC, Bradbury I, et al. Effect of accelerated rehabilitation on function after ankle sprain: randomised controlled trial. BMJ. 10. 2010;340:c1964.

8. Doherty C, Bleakley C, Delahunt E, Holden S. Treatment and prevention of acute and recurrent ankle sprain: an overview of systematic reviews with meta-analysis. Br J Sports Med. 2017;51(2):113–25.

9. Hupperets MDW, Verhagen EALM, Heymans MW, Bosmans JE, van Tulder MW, van Mechelen W. Potential savings of a program to prevent ankle sprain recurrence: economic evaluation of a randomized controlled trial. Am J Sports Med. 2010;38(11):2194–200.

10. Ardèvol J, Bolíbar I, Belda V, Argilaga S. Treatment of complete rupture of the lateral ligaments of the ankle: a randomized clinical trial comparing cast immobilization with functional treatment. Knee Surg Sports Traumatol Arthrosc Off J ESSKA. 2002;10(6):371–7.

11. Calatayud J, Borreani S, Colado JC, Flandez J, Page P, Andersen LL. Exercise and ankle sprain injuries: a comprehensive review. Phys Sportsmed. 2014;42(1):88–93.

12. Hupperets MDW, Verhagen EALM, van Mechelen W. Effect of unsupervised home based proprioceptive training on recurrences of ankle sprain: randomised controlled trial. BMJ. 9. 2009;339:b2684.

13. Verhagen E, Bolling C. Protecting the health of the @hlete: how online technology may aid our common goal to prevent injury and illness in sport. Br J Sports Med. 2015;49(18):1174–8.

14. Wong SJ, Robertson GA, Connor KL, Brady RR, Wood AM. Smartphone apps for orthopaedic sports medicine - a smart move? BMC Sports Sci Med Rehabil. 2015;7:23.

15. Subhi Y, Bube SH, Rolskov Bojsen S, Skou Thomsen AS, Konge L. Expert Involvement and Adherence to Medical Evidence in Medical Mobile Phone Apps: A Systematic Review. JMIR MHealth UHealth. 2015;3(3):e79.

16. van Mechelen DM, van Mechelen W, Verhagen Ealm. Sports injury prevention in your pocket?! Prevention apps assessed against the available scientific evidence: a review. Br J Sports Med. 2014;48(11):878–82.

17. Bondaronek P, Alkhaldi G, Slee A, Hamilton FL, Murray E. Quality of Publicly Available Physical Activity Apps: Review and Content Analysis. JMIR MHealth UHealth. 2018;6(3):e53.

18. Badawy SM, Kuhns LM. Texting and Mobile Phone App Interventions for Improving Adherence to Preventive Behavior in Adolescents: A Systematic Review. JMIR MHealth UHealth. 2017;5(4):e50.

19. Halcomb E, Hickman L. Mixed methods research. Nurs Stand R Coll Nurs. 2015;29(32):41–7.

20. Palinkas LA, Horwitz SM, Green CA, Wisdom JP, Duan N, Hoagwood K. Purposeful sampling for qualitative data collection and analysis in mixed method implementation research. Adm Policy Ment Health. 2015;42(5):533–44.

21. O’Cathain A, Murphy E, Nicholl J. Three techniques for integrating data in mixed methods studies. BMJ. 2010;341:c4587.

22. Chan A-W, Tetzlaff JM, Altman DG, Laupacis A, Gøtzsche PC, Krleža-Jerić K, et al. SPIRIT 2013 statement: defining standard protocol items for clinical trials. Ann Intern Med. 2013;158(3):200–7.

23. Bandholm T, Christensen R, Thorborg K, Treweek S, Henriksen M. Preparing for what the reporting checklists will not tell you: the PREPARE Trial guide for planning clinical research to avoid research waste. Br J Sports Med. 2017;51(20):1494–501.

24. O’Cathain A, Murphy E, Nicholl J. The quality of mixed methods studies in health services research. J Health Serv Res Policy. 2008;13(2):92–8.

25. Castleberry A, Nolen A. Thematic analysis of qualitative research data: Is it as easy as it sounds? Curr Pharm Teach Learn. 2018;10(6):807–15.

26. Julious SA. Issues with number needed to treat. Stat Med. 2005;24(20):3233–5.

27. Guetterman TC, Fetters MD, Creswell JW. Integrating Quantitative and Qualitative Results in Health Science Mixed Methods Research Through Joint Displays. Ann Fam Med. 2015;13(6):554–61.

28. DS93: Luksation og distorsion af led og ligamenter i ankel og fod. Search period: July 3rd, 2018 to April 3rd, 2019. [Internet]. The Danish National Patient Registry. 2017. Available from: https://www.esundhed.dk/Registre/Landspatientsregisteret

29. Vriend I, Coehoorn I, Verhagen E. Implementation of an app-based neuromuscular training programme to prevent ankle sprains: a process evaluation using the RE-AIM Framework. Br J Sports Med. 2015;49(7):484–8.

30. Zheng P, Leff G, Ith MA, Smuck M. App Development for Therapeutic Exercise. PM R. 2017;9(5S):S116–7.

31. Van Reijen M, Vriend I, van Mechelen W, Verhagen EA. Preventing recurrent ankle sprains: Is the use of an App more cost-effective than a printed Booklet? Results of a RCT. Scand J Med Sci Sports. 2018;28(2):641–8.

32. Bleakley C. Supervised physiotherapy for mild or moderate ankle sprain. BMJ. 2016;355:i5984.

33. Brison RJ, Day AG, Pelland L, Pickett W, Johnson AP, Aiken A, et al. Effect of early supervised physiotherapy on recovery from acute ankle sprain: randomised controlled trial. BMJ. 2016;355:i5650.

34. Malliaropoulos N, Ntessalen M, Papacostas E, Longo UG, Maffulli N. Reinjury after acute lateral ankle sprains in elite track and field athletes. Am J Sports Med. 2009;37(9):1755–61.

35. McPhail SM, Schippers M, Marshall AL, Waite M, Kuipers P. Perceived barriers and facilitators to increasing physical activity among people with musculoskeletal disorders: a qualitative investigation to inform intervention development. Clin Interv Aging. 2014;9:2113–22.

36. Liao G-Y, Chien Y-T, Chen Y-J, Hsiung H-F, Chen H-J, Hsieh M-H, et al. What to Build for Middle-Agers to Come? Attractive and Necessary Functions of Exercise-Promotion Mobile Phone Apps: A Cross-Sectional Study. JMIR MHealth UHealth. 2017;5(5):e65.

37. Stevens A, Beurskens A, Köke A, van der Weijden T. The use of patient-specific measurement instruments in the process of goal-setting: a systematic review of available instruments and their feasibility. Clin Rehabil. 2013;27(11):1005–19.

38. Wade DT. Goal setting in rehabilitation: an overview of what, why and how. Clin Rehabil. 2009;23(4):291–5.

39. Horn KK, Jennings S, Richardson G, Vliet DV, Hefford C, Abbott JH. The patient-specific functional scale: psychometrics, clinimetrics, and application as a clinical outcome measure. J Orthop Sports Phys Ther. 2012;42(1):30–42.

40. Picorelli AMA, Pereira LSM, Pereira DS, Felício D, Sherrington C. Adherence to exercise programs for older people is influenced by program characteristics and personal factors: a systematic review. J Physiother. 2014;60(3):151–6.

