## Supplementary Material for "Using the app “Injurymap©” to provide exercise rehabilitation for people with acute lateral ankle sprains seen at the Hospital Emergency Department – a mixed-method pilot study"

### Supplementary A: Good Reporting of A Mixed Methods Study (GRAMMS) checklist

| **Item** | **Reported** |
|---|---|
| Describe the justification for using a mixed methods approach to the research question | Purpose statement, page 3. |
| Describe the design in terms of the purpose, priority, and sequence of methods | Study design, page 4 + Figure 1 |
| Describe each method in terms of sampling, data collection and analysis | Outcomes, pages 6-8 |
| Describe where integration has occurred, how it has occurred and who has participated in it | Study design, page 4 + Figure 1 + Table 5. |
| Describe any limitation of one method associated with the present of the other method | Strengths and limitations, pages 13-14. |
| Describe any insights gained from mixing or integrating methods | Discussion, pages 11-14 |

Reference: O'Cathain A, Murphy E, Nicholl J. The quality of mixed methods studies in health services research. J Health Serv Res Policy. 2008;13(2):92-98.

### Supplementary B: Description of the exercise program.

**Reading guideline:** This exercise program is reported according to the Consensus on Exercise Reporting Template (CERT) [1].

**Materials:** circular rubber band, chair, table, stair step, balance board or pillow.

**Provider:** The exercises are available in the app “InjuryMap”. The program has been developed by two rheumatologists and reviewed by two physiotherapists. All experts had experience in treating ALAS patients. The exercise program was then compared and adjusted to current evidence in exercise rehabilitation for acute ankle sprains.

**Delivery:** It is possible to perform the exercises unsupervised at home. However, participants are free to perform the exercises anywhere they see fit. Participants are encouraged to adhere to the program by reminder notifications on their phone and by the exercise progression. The progression was designed so that participants need to “complete” a number of sessions to be able to progress to more challenging exercise. This resembles some game types and might motivate participant in completing exercise sessions. Data of exercise completion are saved in the app and used to analyze adherence. With each exercise the participants must answer pain level and difficulty. If the participants record no or low pain and low difficulty in an exercise, the app chooses a progression of the exercise next time the participants begin a training session.

**Description:** The exercise program consists of three phases with increasing difficulty. **Phase 1** focuses on stimulating ankle mobility without provoking the acute injury. The exercises are low load with respect for end range motions. **Phase 2** focuses on increasing balance and ankle stability. The phase includes several weight bearing exercises but no impacts. **Phase 3** focuses on strength and mobility. The goal is to return participants to normal activity level. The phase includes strength exercises and jumping exercises with change of direction.

Each phase consists of several exercise categories. The categories are 1) mobility, 2) stability/balance, 3) strength and 4) stretching. Within each phase there can be more than one exercise in each category. An exercise session will consist of minimum one exercise from each category, but not necessarily all exercises in a category.  Exercises can also contain a number of difficulty levels. All exercises in each category must be completed on their highest difficulty before a participant can progress to the next phase. The exercises can potentially cause some discomforts or pain and participants are advised that increased swelling after training session or increased pain until next day should not be tolerated.

A detailed description of the specific exercises in each phase can be found in Tables S1,S2 and S3.

**Table S1: Exercises in phase 1 of the exercise program**

| Phase 1 | | | |
|---|---|---|---|
| **Mobility** | **Stability/balance** | **Strength** | **Stretch** |
| **Mob. 1.1: Ankle bendings.**<br><br>medRxiv policy does not allow for publication of images that may be identifiable to the individual, their friends, family, or neighbors.<br><br>Pictures are available from the corresponding author and from the peer-reviewed paper, once published. | **Stab. 1.1 Ankle Balance**<br><br>medRxiv policy does not allow for publication of images that may be identifiable to the individual, their friends, family, or neighbors.<br><br>Pictures are available from the corresponding author and from the peer-reviewed paper, once published. | **Strength 1.1: Sitting ankle extensions I**<br><br>medRxiv policy does not allow for publication of images that may be identifiable to the individual, their friends, family, or neighbors.<br><br>Pictures are available from the corresponding author and from the peer-reviewed paper, once published. | **Stretch 1.1: Straight leg calf stretch**<br><br>medRxiv policy does not allow for publication of images that may be identifiable to the individual, their friends, family, or neighbors.<br><br>Pictures are available from the corresponding author and from the peer-reviewed paper, once published. |
| Sit on a chair. Lift the one foot from the floor. Bend the foot upwards and downwards as far as possible. If the foot is swelled the exercise can be done lying on the floor with the leg and foot raised above the heart.<br><br>10 reps each foot. | **Level I**<br><br>Stand with shoulder width stance and the feet pointing forward. Bend your knees as far beyond the toes as possible so that the ankles bend to their maximum. You should feel it tightens in the back of the heel and clamps in the front, but it must not be painful. Keep the tempo slow and controlled.<br><br>10 reps. | **Level I:** Sit on a chair with the knees bend. Place the rubber band under one foot just behind the toes. Hold the other end of the rubber band with one hand and tighten it.<br><br>Extend the foot downwards while using the rubber band to manage resistance. Return slowly. Move as far as possible.<br><br>The rubber band needs to be so tight that it is only possible to perform | Stand on a stair step with one foot only touching the step with the forefoot and the heel free from the edge.<br><br>Lower the heel downwards with the knee extended until you feel a stretch in the calf muscles. Put as much weight on the leg as possible without provoking pain.<br><br>Keep stretch position 30 sec. x 3 reps. |

| | | | |
|---|---|---|---|
| | **Level II:** Perform the exercise while looking from side to side. | 10 reps x 3 sets.<br><br>**Level II:** Perform the exercise with the knees extended.<br><br>15 reps x 3 sets. | |
| **Mob. 1.2: Ankle side tilts**<br><br>medRxiv policy does not allow for publication of images that may be identifiable to the individual, their friends, family, or neighbors.<br><br>Pictures are available from the corresponding author and from the peer-reviewed paper, once published. | **Stab. 1.2: Calf raises I**<br><br>medRxiv policy does not allow for publication of images that may be identifiable to the individual, their friends, family, or neighbors.<br><br>Pictures are available from the corresponding author and from the peer-reviewed paper, once published. | **Strength 1.2 Lying ankle bends**<br><br>medRxiv policy does not allow for publication of images that may be identifiable to the individual, their friends, family, or neighbors.<br><br>Pictures are available from the corresponding author and from the peer-reviewed paper, once published. | **Stretch 1.2 Bend knee Calf stretch.**<br><br>medRxiv policy does not allow for publication of images that may be identifiable to the individual, their friends, family, or neighbors.<br><br>Pictures are available from the corresponding author and from the peer-reviewed paper, once published. |
| Sit on a chair. Lift the one foot from the floor. Tilt the foot outwards (eversion) and inwards (inversion) as far as possible<br><br>10 reps each foot. | **Level I:** Stand with shoulder width stance and the feet pointing forward. Raise the heels from the floor so that you're on your tiptoes and return slowly.<br><br>10 reps.<br><br>**Level II:** Perform the exercise while looking from side to side. | Secure the rubber band on a radiator pipe or similar at ground level. Lie on your back with the injured leg extended. Place the end of the rubber band over the back of the foot just behind the toes. Bend the foot upwards while using the rubber band to manage resistance.<br><br>The rubber band needs to be so tight that it is only possible to perform 10 reps for 3 sets. | Stand on a stair step with one foot only touching the step with the forefoot and the heel free from the edge.<br><br>Lower the heel downwards with the knee bended until you feel a stretch in the calf muscles. Put as much weight on the leg as possible without provoking pain.<br><br>Keep stretch position 30 sec. x 3 reps. |

| | | | |
|---|---|---|---|
| **Mob. 1.3: Ankle circles**<br>medRxiv policy does not allow for publication of images that may be identifiable to the individual, their friends, family, or neighbors.<br>Pictures are available from the corresponding author and from the peer-reviewed paper, once published. | **Stab. 1.3: Split stance knee bends I**<br>medRxiv policy does not allow for publication of images that may be identifiable to the individual, their friends, family, or neighbors.<br>Pictures are available from the corresponding author and from the peer-reviewed paper, once published. | **No further progression**<br>medRxiv policy does not allow for publication of images that may be identifiable to the individual, their friends, family, or neighbors.<br>Pictures are available from the corresponding author and from the peer-reviewed paper, once published. | **Stretch 1.3: Hamstring stretch**<br>medRxiv policy does not allow for publication of images that may be identifiable to the individual, their friends, family, or neighbors.<br>Pictures are available from the corresponding author and from the peer-reviewed paper, once published. |
| Sit on a chair. Lift the one foot from the floor. Turn the foot in large circles. Start with 5 rotations in a clockwise direction and then 5 rotations in the opposite direction.<br><br>Repeat twice | **Level I:** Stand with the feet in line with the injured foot in front.<br>Keep the balance while you slowly bend the knees beyond the toes as far as possible, so that the ankle bends to its maximum. You should feel it tightens in the back of the heel and clamps in the front, but it must not be painful.<br>10 repetitions.<br>**Level II:** Perform the exercise while looking from side to side. | | While standing towards a chair put one heel upon the seat. Lower the upper body towards the elevated leg until it stretches in the hamstrings. Be careful not to hyperextend the knees.<br><br>Keep stretch position 30 sec. x 3 reps. |
| **No further progression**<br>medRxiv policy does not allow for publication of images that may be | **Stab. 1.4: Calf raise with knee bend I**<br>medRxiv policy does not allow for publication of images that may be | | **No further progression** |

| | |
|---|---|
| identifiable to the individual, their friends, family, or neighbors.<br><br>Pictures are available from the corresponding author and from the peer-reviewed paper, once published. | identifiable to the individual, their friends, family, or neighbors.<br><br>Pictures are available from the corresponding author and from the peer-reviewed paper, once published. |
| | **Leve I:** Stand with shoulder width stance and the feet pointing forward. Bend your knees as far beyond the toes as possible so that the ankles bend to their maximum. You should feel it tightens in the back of the heel and clamps in the front, but it must not be painful. While bending your knees raise your heels so that you're on your tiptoes and return slowly.<br><br>Repeat 5 times<br><br>**Level II:** Perform the exercise while looking from side to side. |

**Table S2: Exercises in phase 2 of the exercise program**

| Phase 2 | | | |
|---|---|---|---|
| **Mobility** | **Stability/balance** | **Strength** | **Stretch** |
| **Mob. 2.1: Ankle circles**<br><br>medRxiv policy does not allow for publication of images that may be identifiable to the individual, their friends, family, or neighbors.<br><br>Pictures are available from the corresponding author and from the peer-reviewed paper, once published. | **Stab 2.1: One leg balance I**<br><br>medRxiv policy does not allow for publication of images that may be identifiable to the individual, their friends, family, or neighbors.<br><br>Pictures are available from the corresponding author and from the peer-reviewed paper, once published. | **Strength 2.1: Sitting ankle inwards tilt**<br><br>medRxiv policy does not allow for publication of images that may be identifiable to the individual, their friends, family, or neighbors.<br><br>Pictures are available from the corresponding author and from the peer-reviewed paper, once published. | **Stretch 2.1: Straight leg calf stretch**<br><br>medRxiv policy does not allow for publication of images that may be identifiable to the individual, their friends, family, or neighbors.<br><br>Pictures are available from the corresponding author and from the peer-reviewed paper, once published. |
| Sit on a chair. Lift the one foot from the floor. Turn the foot in large circles. Start with 5 rotations in a clockwise direction and then 5 rotations in the opposite direction.<br><br>Repeat twice | **Level I:** From a normal standing position on two legs, gradually place more weight on one leg and lift the other leg from the floor.<br><br>Bend the standing knee slightly and hold for 10 sec<br><br>3 reps. On each leg<br><br>**Level II:** Hold the balance for 20 sec. | Sit on a chair with bended knees and a table leg beside you. Secure the rubber band on the table leg and place the other end on the forefoot just behind the toes. Slowly tilt your foot inwards and return. The heel should be kept on the floor through the movement. Tilt as much in both directions as possible. The knees and hip are kept fixed in the exercise.<br><br>The rubber band needs to be so tight that it is only possible to perform 15 reps for 3 sets. | Stand on a stair step with one foot only touching the step with the forefoot and the heel free from the edge.<br><br>Lower the heel downwards with the knee extended until you feel a stretch in the calf muscles. Put as much weight on the leg as possible without provoking pain.<br><br>Keep stretch position 30 sec. x 3 reps. |

| | | | |
|---|---|---|---|
| | **Level III:** Hold the balance for 10 seconds and look from side to side. | Complete the exercise with both legs | |
| **No further progression** | **Stab. 2.2: Calf raise on one leg**<br><br>medRxiv policy does not allow for publication of images that may be identifiable to the individual, their friends, family, or neighbors.<br><br>Pictures are available from the corresponding author and from the peer-reviewed paper, once published. | **Strength 2.2: Sitting ankle outwards tilt**<br><br>medRxiv policy does not allow for publication of images that may be identifiable to the individual, their friends, family, or neighbors.<br><br>Pictures are available from the corresponding author and from the peer-reviewed paper, once published. | **Stretch 2.2 Bend knee Calf stretch.**<br><br>medRxiv policy does not allow for publication of images that may be identifiable to the individual, their friends, family, or neighbors.<br><br>Pictures are available from the corresponding author and from the peer-reviewed paper, once published. |
| | **Level I:** From a normal standing position on two legs, gradually place more weight on one leg and lift the other leg.<br><br>While standing on one leg raise your heels so that you're on your tiptoes. Hold the balance for a couple of seconds and return slowly.<br><br>Repeat 10 times<br><br>**Level II:** Perform the exercise while looking from side to side. | Sit on a chair with bended knees. Place the rubber band around both feet just behind the toes. Keep the unscathed foot steady while you slowly tilt the injured foot outwards and return. The heel should be kept on the floor through the movement. Tilt as much in both directions as possible. The knees and hip are kept fixed in the exercise.<br><br>The rubber band needs to be so tight that it is only possible to perform 15 reps for 3 sets. | Stand on a stair step with one foot only touching the step with the forefoot and the heel free from the edge.<br><br>Lower the heel downwards with the knee bended until you feel a stretch in the calf muscles. Put as much weight on the leg as possible without provoking pain.<br><br>Keep stretch position 30 sec. x 3 reps. |

| | | | |
|---|---|---|---|
| | | Complete the exercise with both legs | |
| | **Stab. 2.3: Calf raise with knee bend.**<br><br>medRxiv policy does not allow for publication of images that may be identifiable to the individual, their friends, family, or neighbors.<br><br>Pictures are available from the corresponding author and from the peer-reviewed paper, once published. | **No further progression** | **Stretch 2.3: Hamstring stretch**<br><br>medRxiv policy does not allow for publication of images that may be identifiable to the individual, their friends, family, or neighbors.<br><br>Pictures are available from the corresponding author and from the peer-reviewed paper, once published. |
| | **Level I**: From a normal standing position on two legs, gradually place more weight on the injured leg and lift the other leg. Move the unscathed foot behind the injured and slightly put some weight on the toes for balance. From this position bend the knee beyond the toes on the injured leg. While bending, raise your heel from the floor and slowly return. Complete the specified number of repetitions before switching to the other leg. To increase difficulty, look from side to | | While standing towards a chair put one heel upon the seat. Lower the upper body towards the elevated leg until it stretches in the hamstrings. Be careful not to hyperextend the knees.<br><br>Keep stretch position 30 sec. x 3 reps. |

| | | | |
|---|---|---|---|
| | side while performing the repetitions.<br><br>5 reps on each leg<br><br>**Level II**: 7 reps on each leg | | |
| | **No further progression** | | **Stretch 2.4: Bend knee hamstring stretch.**<br><br>medRxiv policy does not allow for publication of images that may be identifiable to the individual, their friends, family, or neighbors.<br><br>Pictures are available from the corresponding author and from the peer-reviewed paper, once published. |
| | | | While standing towards a chair put one heel upon the seat. Bend the elevated knee slightly while lowering the upper body towards the knee until it stretches in the hamstrings. You can place your arms under the thigh for better control of the bended knee.<br><br>Keep stretch position 30 sec. x 3 reps. |

**Table S3: Exercises in phase 2 of the exercise program**

| Phase 3 | | | |
|---|---|---|---|
| Mobility | **Stability/balance** | **Strength** | **Stretch** |
| **Mob. 2.1: Ankle circles**<br>medRxiv policy does not allow for publication of images that may be identifiable to the individual, their friends, family, or neighbors.<br>Pictures are available from the corresponding author and from the peer-reviewed paper, once published. | **Stab. 3.1: Balance on uneven surface**<br>**Level I** **Level II**<br>medRxiv policy does not allow for publication of images that may be identifiable to the individual, their friends, family, or neighbors.<br>Pictures are available from the corresponding author and from the peer-reviewed paper, once published. | **Strength 3.1: Sitting ankle outwards tilt**<br>medRxiv policy does not allow for publication of images that may be identifiable to the individual, their friends, family, or neighbors.<br>Pictures are available from the corresponding author and from the peer-reviewed paper, once published. | **Stretch 3.1: Straight leg calf stretch**<br>medRxiv policy does not allow for publication of images that may be identifiable to the individual, their friends, family, or neighbors.<br>Pictures are available from the corresponding author and from the peer-reviewed paper, once published. |
| Sit on a chair. Lift the one foot from the floor. Turn the foot in large circles. Start with 5 rotations in a clockwise direction and then 5 rotations in the opposite direction.<br>Repeat twice | **Level I:** With a slightly bended knee stand on one leg on a balance board, hard pillow (like a couch pillow) or similar. Hold the balance without the other leg touches the ground. Repeat with the other leg.<br>Hold balance for 1 min. on each leg.<br>**Level II:** With slightly bended knees stand on both legs on a balance board, hard | Sit on a chair with bended knees. Place the rubber band around both feet just behind the toes. Keep the unscathed foot steady while you slowly tilt the injured foot outwards and return. The heel should be kept on the floor through the movement. Tilt as much in both directions as possible. The knees and hip are kept fixed in the exercise.<br>The rubber band needs to be so tight that it is only possible to perform 15 reps for 3 sets. | Stand on a stair step with one foot only touching the step with the forefoot and the heel free from the edge.<br>Lower the heel downwards with the knee extended until you feel a stretch in the calf muscles. Put as much weight on the leg as possible without provoking pain.<br>Keep stretch position 30 sec. x 3 reps. |

| | | | |
|---|---|---|---|
| | pillow (like a couch pillow) or similar. Slowly and with control shift your weight back on your heels and forth on your toes for 20 sec. Afterwards shift your weight from side to side for 20 sec.<br><br>Hold the balance in total for 1 min. | Complete the exercise with both legs | |
| | **Stab. 3.2: One leg balance II**<br><br>medRxiv policy does not allow for publication of images that may be identifiable to the individual, their friends, family, or neighbors.<br><br>Pictures are available from the corresponding author and from the peer-reviewed paper, once published. | **Strength 3.2: Squat**<br><br>medRxiv policy does not allow for publication of images that may be identifiable to the individual, their friends, family, or neighbors.<br><br>Pictures are available from the corresponding author and from the peer-reviewed paper, once published. | **Stretch 3.2: Bend knee Calf stretch**<br><br>medRxiv policy does not allow for publication of images that may be identifiable to the individual, their friends, family, or neighbors.<br><br>Pictures are available from the corresponding author and from the peer-reviewed paper, once published. |
| | From a normal standing position on two legs, gradually place more weight on one leg and lift the other leg from the floor.<br><br>Bend the standing knee slightly and hold for 10 sec while looking from side to side | Place a chair a hands length behind you and stand with shoulder width stance. Weight should be equally distributed on both legs.<br><br>Bend your knees as would you sit down on the chair but halt the movement just before you are sitting. Slowly return to upright position. Keep the | Stand on a stair step with one foot only touching the step with the forefoot and the heel free from the edge.<br><br>Lower the heel downwards with the knee bended until you feel a stretch in the calf muscles. Put as much weight |

| | | | |
|---|---|---|---|
| | 3 reps. On each leg | knees oriented parallel with feet through the movement.<br><br>15 reps x 3 sets. | on the leg as possible without provoking pain.<br><br>Keep stretch position 30 sec. x 3 reps. |
| | | **Strength 3.3: Lounges**<br><br>medRxiv policy does not allow for publication of images that may be identifiable to the individual, their friends, family, or neighbors.<br><br>Pictures are available from the corresponding author and from the peer-reviewed paper, once published. | **Stretch 3.3 Hamstring stretch**<br><br>medRxiv policy does not allow for publication of images that may be identifiable to the individual, their friends, family, or neighbors.<br><br>Pictures are available from the corresponding author and from the peer-reviewed paper, once published. |
| | | Stand with a shoulder width stance. Make a large step forward and lower your hips until both knees are bent at about a 90-degree angle and return to upright position. Make sure your front knee is directly above your ankle and keep your upper body straight with your shoulders relaxed.<br><br>Repeat 15 times and switch leg. 3 sets for each leg. | While standing towards a chair put one heel upon the seat. Lower the upper body towards the elevated leg until it stretches in the hamstrings. Be careful not to hyperextend the knees.<br><br>Keep stretch position 30 sec. x 3 reps |
| | | **Strength 3.4: Jumps**<br><br>**Level I** **Level II**<br><br>**Level III** **Level IV** | **Stretch 3.4 Bend knee hamstring stretch.**<br><br>medRxiv policy does not allow for publication of images that may be |

| | | | |
|---|---|---|---|
| | | medRxiv policy does not allow for publication of images that may be identifiable to the individual, their friends, family, or neighbors.<br><br>Pictures are available from the corresponding author and from the peer-reviewed paper, once published. | identifiable to the individual, their friends, family, or neighbors.<br><br>Pictures are available from the corresponding author and from the peer-reviewed paper, once published. |
| | | **Level I: Straight jump**<br>Stand with a shoulder width stance. Jump straight upwards and land controlled with equal weight on both legs. Make sure that the knees are oriented over the foot in the landing, so that they do not fall inwards.<br><br>5 reps x 1 set<br><br>**Level II: One leg straight jump**<br>Stand on one leg. Jump straight upwards and land by controlling that the knee are oriented over the foot though the landing. The knee should not fall inwards. It should feel even on both legs.<br><br>5 reps on each leg. | While standing towards a chair put one heel upon the seat. Bend the elevated knee slightly while lowering the upper body towards the knee until it stretches in the hamstrings. You can place your arms under the thigh for better knee control<br><br>Keep stretch position 30 sec. x 3 reps. |

| | | |
|---|---|---|
| | | **Level III: One leg forward jump**<br>Stand on one leg and jump back and forward. Find the balance on each landing before you jump again. It should feel even on both legs.<br><br>5 reps x 2 sets on each leg<br><br>**Level IV: One leg side jump**<br>Stand on one leg and jump from side to side. Find the balance on each landing before you jump again. It should feel even on both legs.<br><br>5 reps x 2 sets on each leg. |

### Supplementary C: Completed exercise sessions per participant.

**Figure S1: Number of completed exercise sessions per participant. EXses = exercise sessions.**

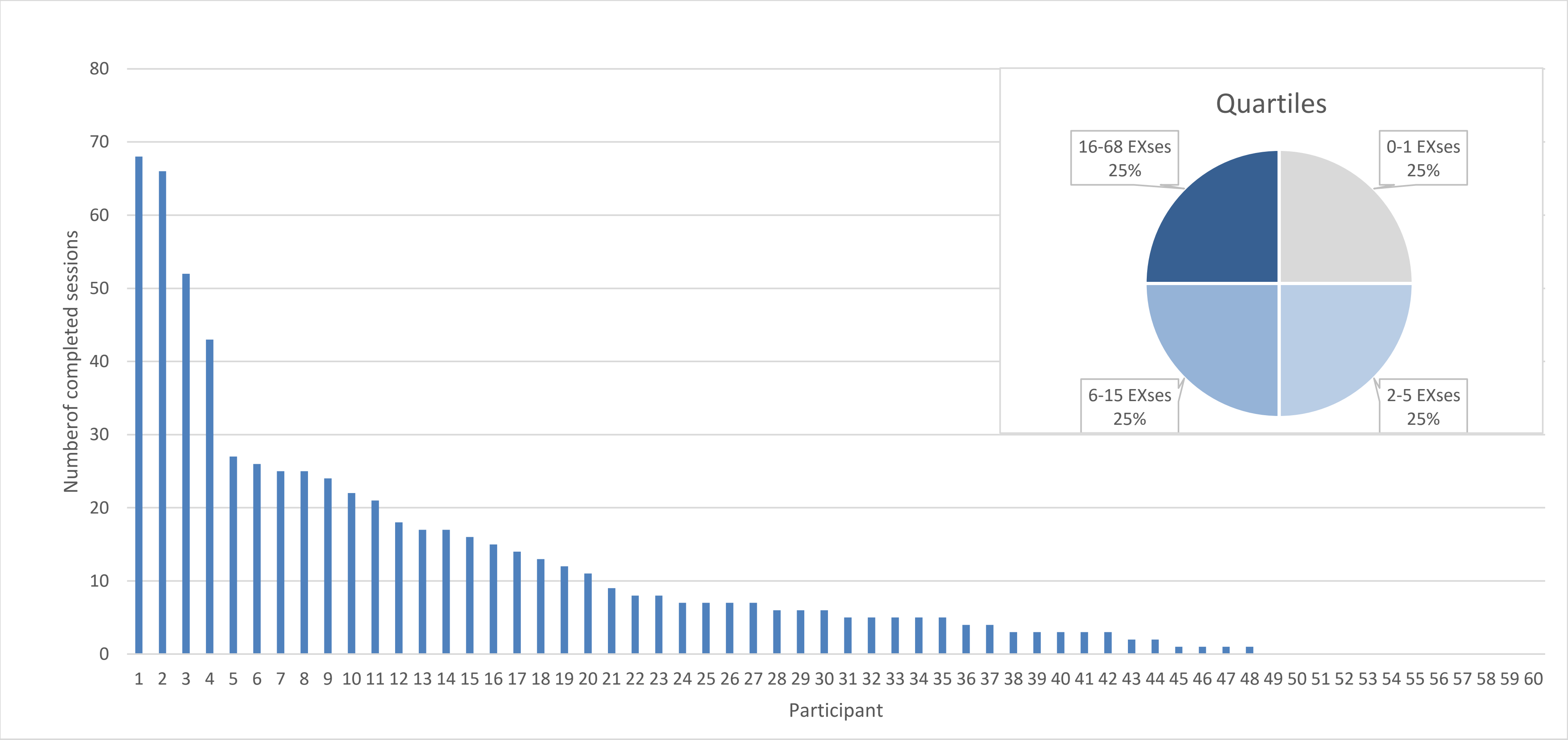

### Supplementary D: Adherence by group.

This exploratory analysis of the adherence by group was conducted after the planned analysis and contains small and different sized groups. Interpretations should be done with caution. Participants with a bachelor degree or higher completed on average 15 exercise sessions, and participants with a shorter education completed on average 8 exercise session. Participants 50 years of age or more completed on average 22 sessions and participants with ages from 10 to 18 yr completed on average 4 sessions.

**Figure S2: Adherence by different grouping variables.**

| Baseline charateristic | Item | N participants | Completed exercise sessions (mean) |
|---|---|---|---|
| *Education level* | Shorter than bachelor | 27 | 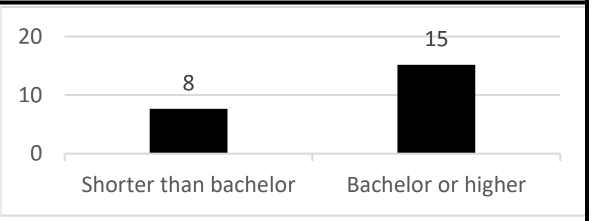 |
| | Bachelor or higher | 29 | |
| *Age group* | 10-18 | 9 | 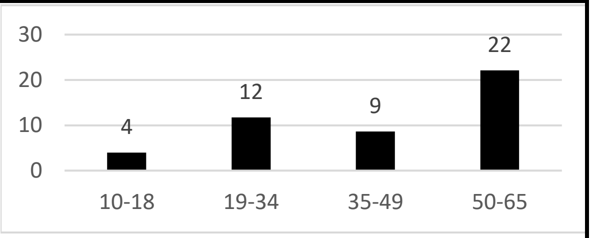 |
| | 19-34 | 24 | |
| | 35-49 | 19 | |
| | 50-65 | 8 | |
| *Sports active* | Sports active | 12 | 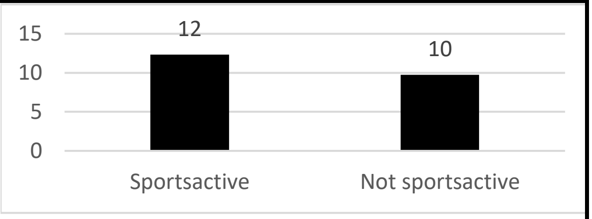 |
| | Not sports active | 10 | |
| *Work demands* | Mostly sitting | 12 | 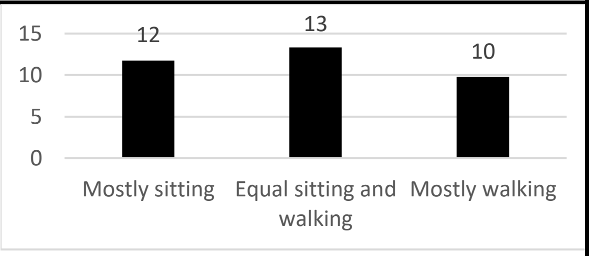 |
| | Equal sitting and walking | 13 | |
| | Mostly walking | 10 | |

### References

1. Slade SC, Dionne CE, Underwood M, Buchbinder R: Consensus on Exercise Reporting Template (CERT): Explanation and Elaboration Statement. *Br J Sports Med* 2016.
